# Rare variants in the outcome of social skills group training for autism

**DOI:** 10.1101/2021.05.19.21257395

**Authors:** Danyang Li, Nora Choque Olsson, Martin Becker, Abishek Arora, Hong Jiao, Nina Norgren, Ulf Jonsson, Sven Bölte, Kristiina Tammimies

**Affiliations:** Center of Neurodevelopmental Disorders (KIND), Centre for Psychiatry Research; Department of Women’s and Children’s Health, Karolinska Institutet and Child and Adolescent Psychiatry, Stockholm Health Care Services, Stockholm County Council, Stockholm, Sweden; Astrid Lindgren Children’s Hospital, Karolinska University Hospital, Region Stockholm, Sweden; Department of Psychology, Stockholm University, Stockholm, Sweden; Center for Psychiatry Research, Department of Clinical Neuroscience, Karolinska Institutet and Stockholm Health Care Services, Stockholm County Council, Stockholm, Sweden; Department of Biosciences and Nutrition, Karolinska Institutet, and Clinical Research Centre, Karolinska University Hospital, Huddinge, Sweden; Department of Molecular Biology, National Bioinformatics Infrastructure Sweden (NBIS), Science for Life Laboratory, Umeå University, 901 87 Umeå, Sweden; Department of Neuroscience, Child and Adolescent Psychiatry, Uppsala University, Sweden; Curtin Autism Research Group, Curtin School of Allied Health, Curtin University, Perth, Western Australia

**Keywords:** autism spectrum disorder, social skills group training, exome sequencing, rare variants

## Abstract

**Background:** Exome sequencing has been proposed as the first-tier genetic testing in autism spectrum disorder (ASD). Here, we performed exome sequencing in autistic individuals with average to high intellectual abilities (N = 207) to identify a molecular diagnosis of ASD and genetic modulators of intervention outcomes following social skills group training (SSGT) or standard care.

**Methods:** Within a randomized controlled trial of SSGT, we performed exome sequencing to prioritize variants of clinical significance (VCSs), variants of uncertain significance (VUSs) and generated a pilot scheme to calculate genetic scores representing the genetic load of rare and common variants in the synaptic transmission pathway (GSSyT_r_ and GSSyT_c_). The association with the primary outcomes (parent-reported autistic traits, Social Responsiveness Scale) was computed using a mixed linear model. Behavioral and genetic features were combined in a machine learning (ML) model to predict the individual response within the cohort.

**Results:** In total, 4.4% (n = 9) and 11.3% (n = 23) of the cohort carried VCSs and VUSs, respectively. Compared to non-carriers, individuals with VCS or VUS together tended to have limited improvements of the interventions (β = 9.22; CI (−0.25, 18.70); P = 0.057) and improved significantly less from standard care (β = 9.35; CI (0.70, 18.00); P = 0.036), but not from SSGT (β = -2.50; CI (−13.34, 8.35); P = 0.65). In addition, both GSSyT_r_ and GSSyT_c_ were associated with differential outcomes in standard care and SSGT groups. Our ML model showed the importance of rare variants for outcome prediction.

**Conclusions:** Autistic individuals with likely pathogenic rare variants identified by exome sequencing might benefit less from the standard care. SSGT could therefore be of heightened importance for this subgroup. Further studies are needed to understand genetic predisposition to intervention outcomes.

**Trial registration:** Social Skills Group Training (“KONTAKT”) for Children and Adolescent With High-functioning Autism Spectrum Disorders (KONTAKT-RCT), NCT01854346. Submitted 20 April 2013 - Retrospectively registered, https://clinicaltrials.gov/ct2/show/record/NCT01854346?view=record

## Background

The molecular genetic understanding of autism spectrum disorder (ASD) has undergone breakthroughs in research settings over the last decade, and these findings are continuously translated into clinical use [1,2]. Currently, the main implication of clinical genetic testing for an individual with ASD is to potentially identify a molecular diagnosis that can be used as a diagnosis specifier according to the Diagnostic & Statistical Manual of Mental Disorders (DSM-5) [3]. The standard recommended genetic testing for individuals with ASD has been targeted testing for Fragile X syndrome and chromosomal microarray to detect copy number variations (CNVs) [4]. However, a recent consensus statement suggested that exome sequencing detecting rare genetic variants should replace chromosomal microarray as the first-tier genetic testing for neurodevelopmental disorders (NDDs), including ASD [5]. Despite these recommendations, the utilization of genetic testing in ASD is still low [6], and the usefulness of the genetic information beyond a molecularly defined diagnosis is still limited. In addition to rare genetic variants, polygenic risk score (PRS), which aggregates the risk of common variants from genome-wide association studies (GWAS), is associated with ASD and shows an additive effect with rare variants [7,8]. A justified assumption is that genetic information can help to develop more individualized and effective interventions [9]. Nevertheless, few studies are focused on investigating how genetic information or genetically defined subgroups of autistic individuals can be used to guide the choice or development of treatments and interventions for ASD [10].

Genetic findings in ASD have shown important insights into the biology of the condition by suggesting the convergence of gene networks such as synaptic transmission, gene regulation, and chromatin remodeling pathways [11–13]. Machine learning (ML) prediction models are increasingly being used in relation to ASD, and studies have successfully used genetic information to predict additional genes, diagnosis, and variants pathogenicity [14–16]. However, a few studies have used ML methods to predict intervention outcomes in ASD and other mental disorders by integrating genetic and clinical predictors [17,18].

With challenges in social communication and interaction being core features of ASD, the most common interventions offered to autistic individuals target social skills or development [19]. Social skills group training (SSGT) is a behavioral intervention widely applied in ASD to ameliorate social communication difficulties, with varied responses depending on factors such as age and sex [20]. In addition, our group demonstrated that both rare CNVs and common variant PRS could affect individual intervention outcomes in a randomized controlled trial evaluating the SSGT KONTAKT® [21,22]. Seeking to examine further the role of rare variants on individual outcomes of SSGT and standard care, we performed exome sequencing in the participants and leveraged all level genetic data and clinical information from the KONTAKT® trial [20], as summarized in Figure 1. Additionally, we proposed a pilot scheme on how to construct a gene set specific genetic score measuring the contribution of common and rare variants in synaptic transmission genes to gain biological insight for outcome variability. Lastly, we built a machine learning model combining clinical and genetic features to predict intervention responses within the KONTAKT® randomized controlled trial.

**Figure 1.**
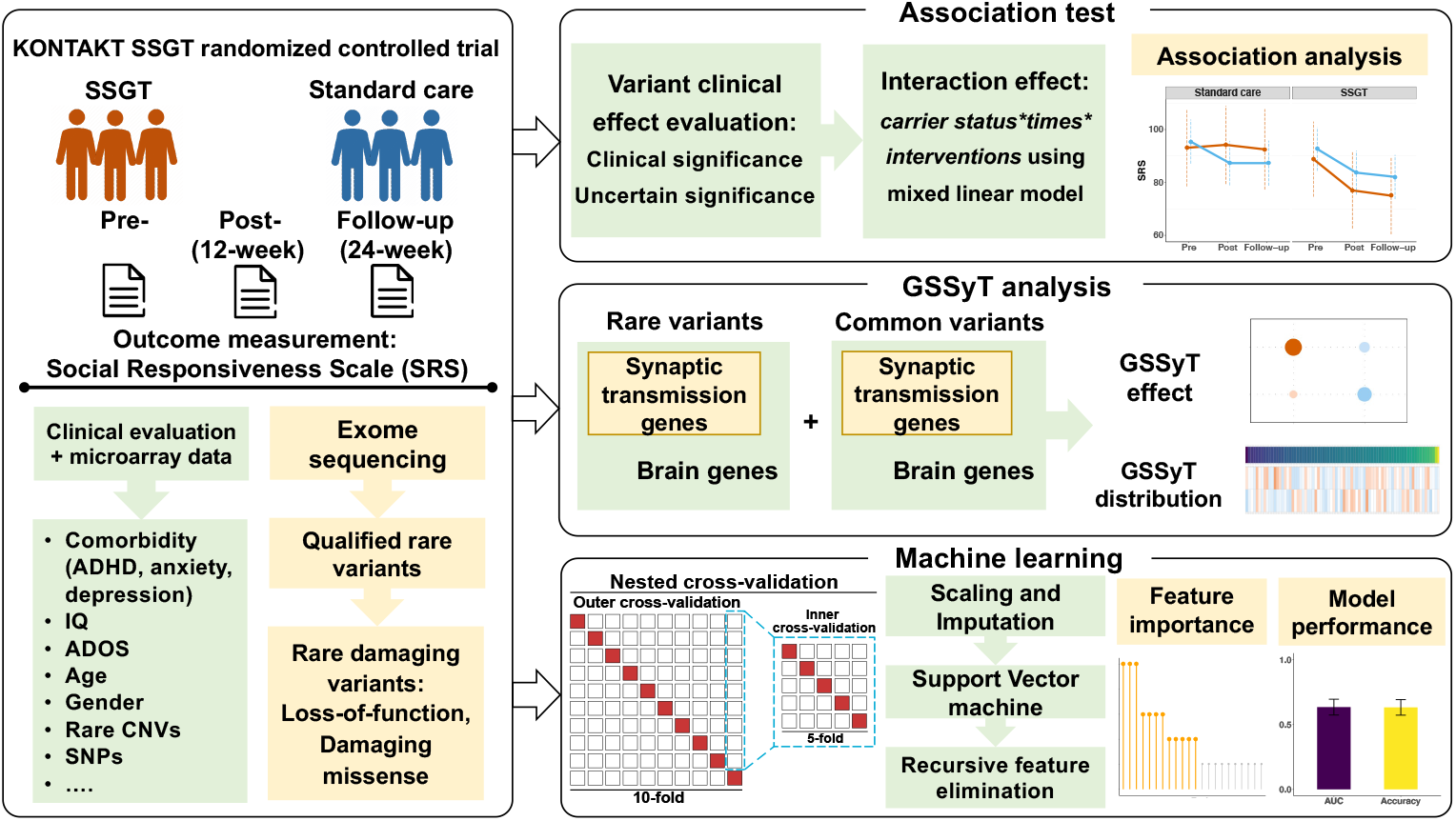
Overview of the study and performed analysis. SSGT: social skills group training; ADHD: attention deficit hyperactivity disorder; ADOS: Autism Diagnostic Observation Schedule; CNV: copy number variation; SNP: single nucleotide polymorphism; GSSyT: genetic score for synaptic transmission genes.

## Methods

### Participants

All participants in this study were a part of the SSGT KONTAKT® randomized controlled trial (clinical trial identifier: NCT01854346) [20]. The inclusion and exclusion criteria, participant characteristics, and the trial outcome have been reported in detail earlier [20]. In brief, all the participants had a confirmed ASD diagnosis, had an IQ > 70, and at least one comorbid neurodevelopmental or psychiatric diagnosis (such as attention deficit hyperactivity disorder (ADHD), anxiety disorder, or depression). In KONTAKT®, participants are trained in groups of children (7-13 years) or adolescents (13-17 years). In the trial, participants were randomly assigned to either the standard care receiving any usually provided clinical intervention, or the SSGT group receiving standard care plus SSGT. A total of 207 (SSGT group: 105, standard care group: 102) participants were included in this study, who had consented to the genetic part and had available saliva samples as well as recorded parent-reported Social Responsiveness Scale (SRS) [23] total scores (primary outcome) at baseline and either at post-intervention (n = 186) or at 12-week follow-up (n = 169).

The four subscales of the SRS probing different social behavioral traits (social awareness, social cognition, social motivation, social communication) were also used in this study as targeted outcomes [23]. As a part of the analysis presented in this study, we determined a binary classification outcome based on the change in the SRS from pre-to post-treatment. Participants with a >10% decrease in the total SRS score were classified as “response to treatment”. Additional information on the other clinical measures can be found in Supplementary Information.

### Variant calling and prioritization

Exome sequencing was done on NovaSeq 6000 using paired-end 2 × 150 readout. Detailed information on library preparation and quality control can be found in Supplementary Information. After sequencing, data quality control, reads alignment, variant calling, and annotations were completed using Mutation Identification Pipeline (MIP) [24] (https://github.com/Clinical-Genomics/MIP). The average sequencing coverage of OMIM genes in all subjects was 77.9, and 98.2% OMIM gene exons reached 10X sequence depth (Supplementary Fig 1). In addition, 89.9% and 88.5% of the exome were covered with at least 10X and 20X sequence depth, respectively.

The called variants were first filtered based on quality and minor allele frequency (Supplementary Information). The variant effect, defined by the Sequence Ontology (http://www.sequenceontology.org/), was annotated by Ensembl Variant Effect Predictor (VEP) v92. After quality control, qualified rare variants with the effect of loss-of-function (LOF, splice donor/acceptor, frameshift, start lost, stop gained) or damaging missense were prioritized as rare deleterious variants for clinical significance assessment. Rare deleterious variants that were located in developmental disorder-related genes (Supplementary Table 1, see Supplementary Information), and in accordance with the gene inheritance pattern were considered developmental disorder-related rare variants. Furthermore, evaluation to categorize the developmental disorder-related rare variants to pathogenic/likely pathogenic, uncertain clinical effect, or benign/likely benign based on ACMG guideline was done [25]. Both pathogenic and likely pathogenic variants were classified as variants of clinical significance (VCSs). All VCSs and variants of uncertain significance (VUSs) were validated by Sanger sequencing.

### Genetic score for synaptic transmission (GSSyT)

To test a pilot scheme for analyzing gene set specific genetic load, we constructed genetic scores for synaptic transmission (SyT) genes (GSSyT). To calculate GSSyT based on the rare genetic variants (GSSyT_r_), we used the number of genes with rare deleterious exome variants in the synaptic transmission gene set and brain gene set combining with rare CNV genes [21] (Supplementary Information). The number of synaptic genes captured by two kinds of rare variants was added and divided by the number of brain-expressed genes with rare variants in each individual. Next, we generated GSSyT for the common variants (GSSyT_c_) acquired from the previous study [22]. The calculation of GSSyT_c_ was based on set-based polygenic risk score (PRS) using a new feature of PRSice named PRSet (https://www.prsice.info/quick_start_prset/). We chose the largest ASD GWAS of Grove et al. [7] as reference data to calculate set-based PRS for ASD. The ratio between synaptic transmission and brain PRSs was represented as GSSyT_c_.

### Statistical analyses

We applied a mixed linear model to assess whether VCS/VUS carrier status was associated with intervention outcome measured by parent-reported SRS total score and its four subscales. We included a three-way interaction: carrier status*time points*interventions together with all lower-order interactions, age, sex, PRS for ADHD (P-threshold 1.0), clinically significant CNVs as fixed factors, and the clinics in which the individuals had participated in the intervention and individual IDs as random factors in the model. In addition, to measure the separate effect of VCS/VUS on SSGT or standard care, the two-way interactions of carrier status*time points were tested using the same mixed linear model in SSGT and standard care subgroups separately. Furthermore, we also performed linear regression as a secondary model to test whether VCS/VUS and ASD PRS (P-threshold 0.5) independently affect the intervention outcomes.

Similarly, we conducted a mixed linear model to investigate the effect of the variants from the synaptic transmission pathway on the intervention outcomes. We combined two three-way interactions of GSSyT_r_*time points*interventions and GSSyT_c_*time points*interventions, together with age, sex as fixed factors, and same random factors as earlier. We also tested the effect of GSSyT_c_ and GSSyT_r_ in two intervention subgroups using the two-way interaction of GSSyT*time points. Furthermore, linear regression was performed to test the interaction of GSSyT_c_ and GSSyT_r_ using the change in SRS total score and sex and age as cofactors.

### Machine learning prediction

As an outcome for our machine learning prediction, a binary outcome for response to treatment (>10% decrease in the total SRS score) or not was calculated. The prediction model was constructed using the participant characteristics, comorbidity diagnoses, treatments, autism-related scales assessed at pre-treatment, and genetic information of rare and common variants. Detailed feature information is listed in Supplementary Table 2. We performed nested cross-validation by splitting total data into training and validation sets using outer cycle (10-fold) and then splitting the training set again into training and test data using inner cycle (5-fold). A linear support vector machine (SVM) was employed for prediction, and recursive feature elimination was implemented to rank the most important features. We used the area under the receiver operating characteristic curve (AUC) as model performance measurement. Other metrics, such as sensitivity, specificity, and accuracy, were evaluated at each outer cycle cross-validation.

More detailed information can be found in Supplementary Information.

## Results

### Clinically significant variants and phenotype characteristics

Exome sequencing data were analyzed for 207 individuals from the SSGT KONTAKT® randomized controlled trial. Three individuals were excluded due to the low sample quality. In the remaining data from 204 individuals, 102 (50.0%) were from the SSGT group, 60 (29.4%) were female, and 124 (60.8%) were adolescents. Based on our variant prioritization scheme, each individual had on average 54 rare deleterious variants, of which four were putatively developmental disorder-related rare variants (Supplementary Fig 2). From these, we identified nine variants of clinical significance (VCSs) in nine carriers (4.4%) and 24 variants of uncertain significance (VUSs) in 23 carriers (11.3%) following ACMG guideline [25] (Table 1, Supplementary Table 3). All VCSs (five indels, four single nucleotide variations (SNVs)) were rare loss-of-function variants in autosomal dominant genes (Table 1). In total, 29 individuals had either one VCS or VUS, and two individuals carried two different variants (one individual with a VUS and a VCS, and another individual with two VUSs) respectively. There were no significant differences in parental reported total Social Responsiveness Scale (SRS) score at pre-treatment (P = 0.68), IQ (P = 0.86), or Autism Diagnostic Observation Schedule (ADOS) social communication total scores (P = 0.28) between individuals with VCS/VUS and those without these variants (Supplementary Fig 3). We also tested the difference after removing individuals with earlier reported clinically significant CNVs [21], and found similar non-significant results (pre-treatment SRS: P = 0.69, IQ: P = 0.82, ADOS: P = 0.30).

**Table 1.**
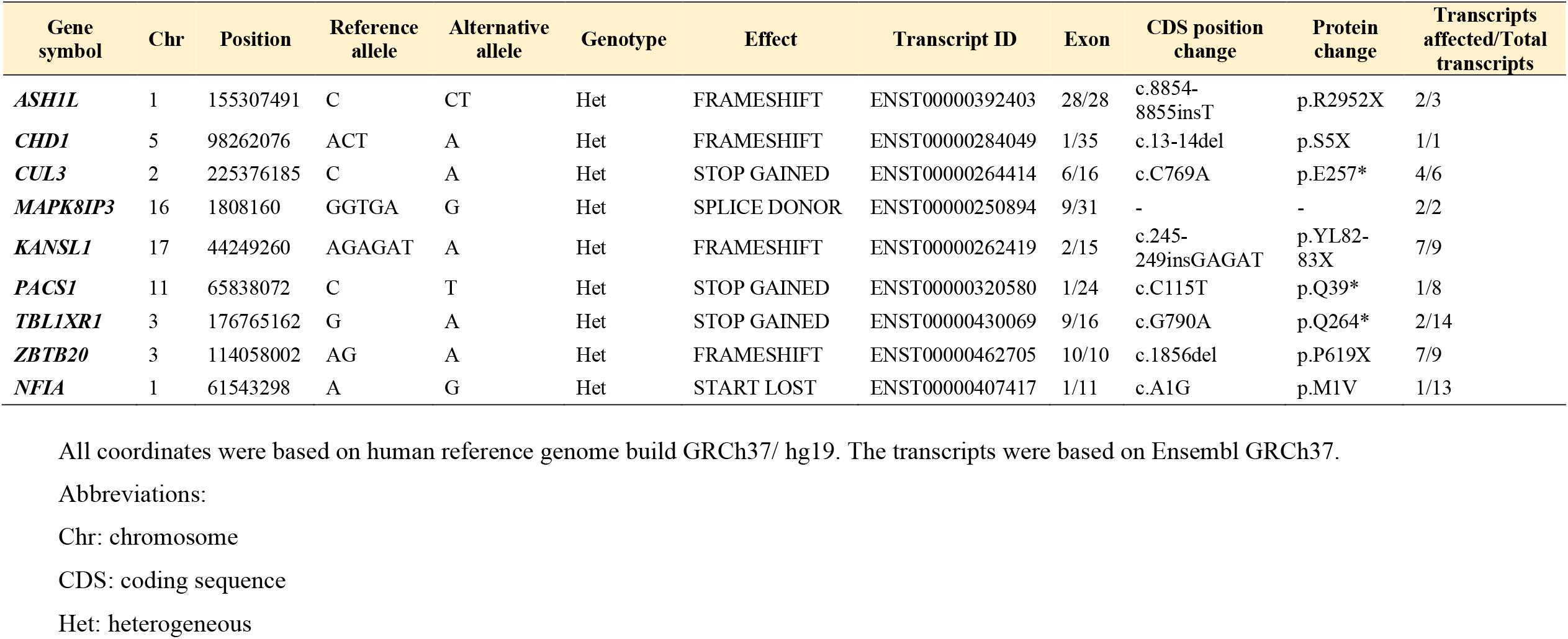
Detailed information of the prioritized clinically significant variants identified in the cohort.

In addition to exome sequencing, we used our earlier acquired data on clinically significant copy number variations (CNVs) and common variants polygenic risk score (PRS) for ASD [21,22]. The distribution of PRS for ASD did not differ between individuals with or without VCS/VUS from exome sequencing (P = 0.37, Supplementary Fig 4a). Two individuals were carriers of both clinically significant CNVs and VUSs, but no individual had both clinically significant CNVs and VCSs (P = 0.65, Supplementary Fig 4b). Detailed distribution of PRS for ASD and carriers with different prioritized variants were shown in Supplementary Fig 4c.

### Association between carrier status and SRS score

To investigate the role of VCS/VUS identified in this study on the SSGT and standard care intervention outcomes, we used parent-reported autistic traits total score on the SRS as the primary outcome and tested interactions of carrier status of prioritized variants, time points, and interventions using a mixed linear model in the whole cohort. We first analyzed individuals with VCS or VUS together in comparison with non-carriers. The interaction of carrier status and time points indicated that individuals who were carriers tended to improve less at post-intervention (β = 9.22, P = 0.057, Supplementary Table 4, Fig 2). Further analysis of the changes in the four SRS subscales (social communication, social awareness, social cognition, social motivation) showed that the total SRS score differences were driven by changes in the social cognition subscale ((VCS + VUS)*post-intervention: β = 3.34, P = 0.006, Supplementary Fig 5). The other three subscales did not show any significant associations with the carrier status (Supplementary Fig 5). A similar trend was observed when comparing carriers of VCS to non-carriers. We found VCS carriers had significantly less improvements at post-intervention (β = 14.84, P = 0.029, Supplementary Table 4), especially for the changes of social cognition subscale (VCS*post-intervention: β = 4.38, P = 0.0095). No outcome difference was detected between VUS carriers and non-carriers (VUS*post-intervention: β = 4.36, P =0.49, Supplementary Table 4). We did not find any significant association at follow-up either (Supplementary Table 4). When contrasting the two intervention groups in three-way interaction, no significant association was observed with SRS total scores or any of the four subscales (Fig 2, Supplementary Table 4, Supplementary Fig 5). Next, to distinguish the influence of the VCS/VUS within each intervention group, we performed the association analysis for the intervention groups separately. When combining VCS and VUS carriers into a single carrier group, significantly worse outcome was found in carriers receiving standard care compared to non-carriers at post-intervention (β = 9.35, P = 0.036, Supplementary Table 5). Similarly, VCS carriers in the standard care group also improved less than non-carriers at post-intervention (β = 14.86, P = 0.017, Supplementary Table 5). No significant association was found in the SSGT group (Supplementary Table 5).

**Figure 2.**
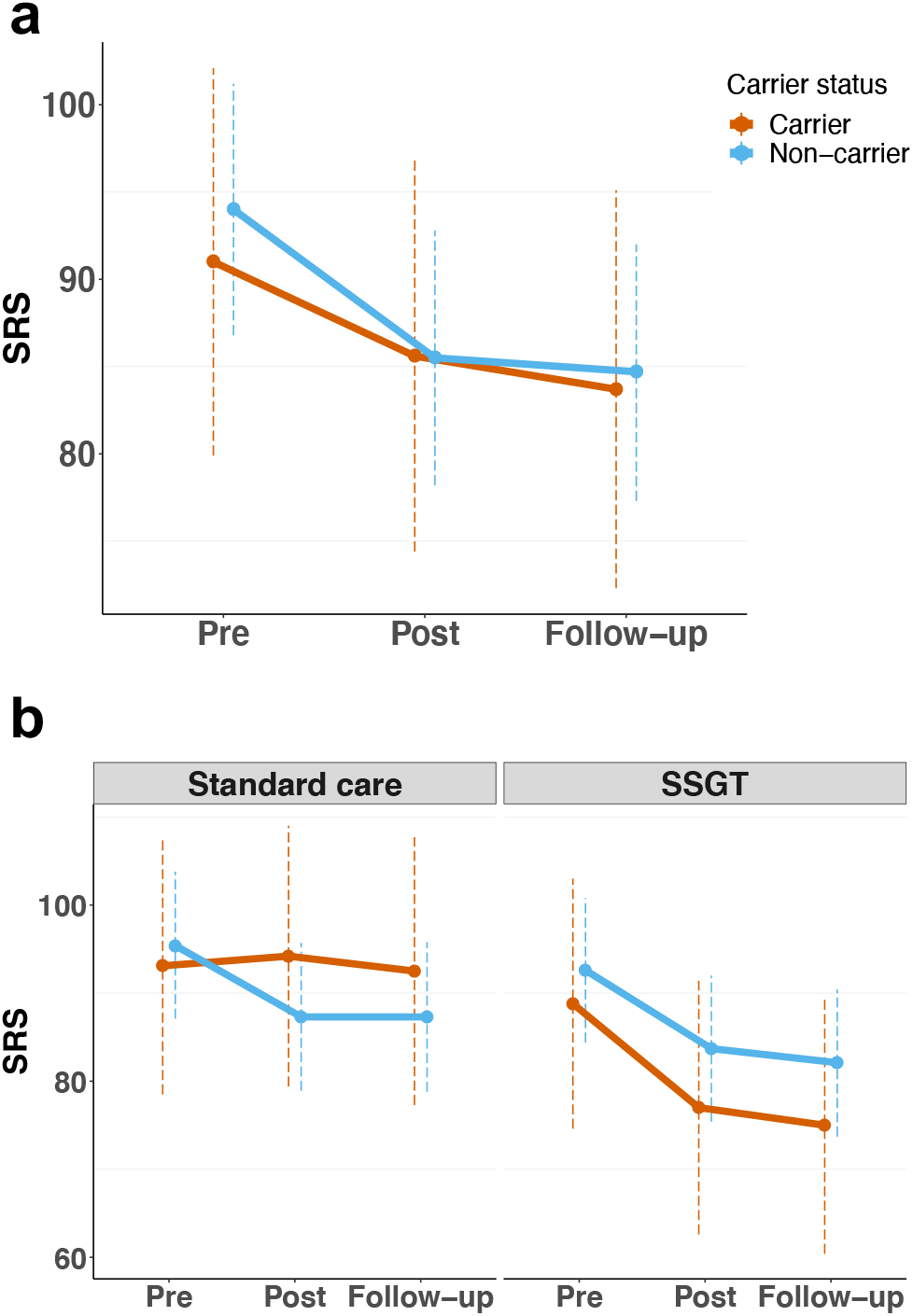
Association between VCS+VUS carrier status and intervention outcomes. **a**. Least-squares means of Social Responsiveness Scale (SRS) among carriers at pre-, post-intervention and follow-up. **b**. Least-square means of SRS among carriers at pre-, post-intervention and follow-up in standard care and social skills group training (SSGT) groups. VCS: variant of clinical significance; VUS: variant of uncertain significance.

We also tested the interaction of PRS for ASD from our previous study [22] and VCS/VUS in the linear model. Outcomes at post-intervention and follow-up were measured by SRS changes at two time points compared to pre-intervention. Similarly, we found carriers of VCS had less improvements at post-intervention (VCS: β = 14.66, P = 0.03). No significant associations were found either in the two-way (exome variant*PRS) or the three-way interactions (exome variant*PRS*interventions), demonstrating the independent effect of PRS and rare pathogenic exome variants on treatment outcomes (Supplementary Fig 6).

### Synaptic transmission gene set on intervention outcomes

Based on our earlier findings of synaptic gene sets and the intervention outcome [22], we generated a pilot scheme to calculate gene set specific scores based on both rare (rare deleterious exome variants and rare CNVs) and common variants from synaptic transmission genes and assessed their effect on interventions. Protein coding genes expressed in the brain were used as a background gene set. We calculated genetic scores in synaptic transmission gene set (GSSyT) for each individual to separately represent the genetic load of rare (GSSyT_r_) and common variants (GSSyT_c_), as shown in Supplementary Fig 7. There was no correlation between the scores generated from the rare and common variants affecting synaptic transmission genes (Pearson r = 0.069, P = 0.35) and no statistical difference of GSSyT_r_ and GSSyT_c_ between two intervention groups (GSSyT_r_: t = 0.65, P = 0.51; GSSyT_c_: t = 0.11, P = 0.91). The GSSyT_c_ had no association with the intervention outcomes in the whole cohort (Supplementary Table 6). However, individuals with higher GSSyT_c_ in the SSGT group had better outcomes compared with the standard care group at post-intervention (β = -5.52, P = 0.033, Fig 3). On the contrary, we found a significant outcome improvement at follow-up in individuals with higher GSSyT_r_ in the whole cohort (β = -5.38, P = 0.016, Supplementary Table 6). Further, individuals with higher GSSyT_r_ had less improvements after SSGT compared with standard care at follow-up (β = 8.30, P = 0.0044, Fig 3). After testing GSSyT effect in each intervention group, we found similar associations where higher GSSyT_r_ was beneficial for outcome within the standard care (β = -5.46, P = 0.0091, Supplementary Table 7), and higher GSSyT_c_ was associated with favorable outcome after SSGT (β = -5.42, P = 0.0062, Supplementary Table 7).

**Figure 3.**
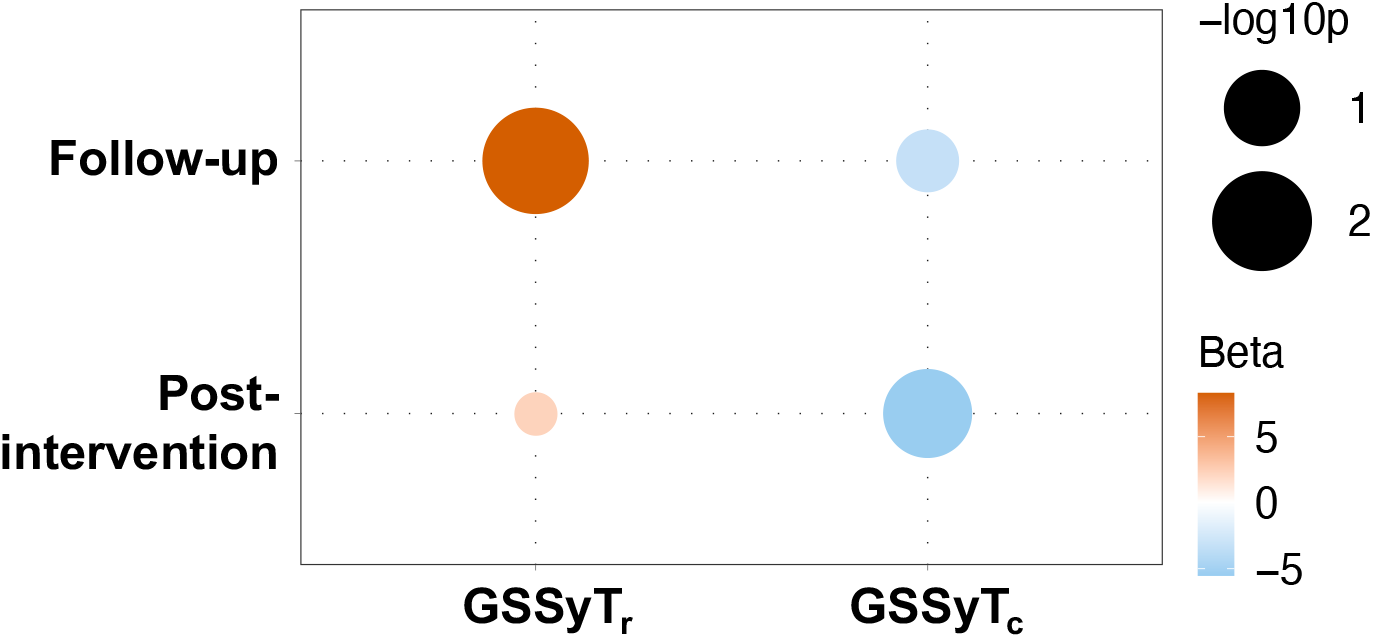
The effect of rare and common variant genetic scores for synaptic transmission genes (GSSyT_r_ and GSSyT_c_) on intervention outcomes at post-intervention and follow-up. The color of each circle represents the coefficient value in 3-way interaction of GSSyT_r_/GSSyT_c_*time points*interventions. The size of each circle is the coefficient -log10p value calculated from the mixed linear model.

To further test the relationship between GSSyT_r_ and GSSyT_c_, we also used linear regression adding the interaction effect of GSSyT_r_ and GSSyT_c_ together with interventions using SRS changes of post-intervention and follow-up. Similar to previous results, improved effects of both GSSyT_r_ and GSSyT_c_ were indicated on interventions (post-intervention: GSSyT_c_*SSGT: β = -5.95, P = 0.028; follow-up: GSSyT_r_: β = -6.67, P = 0.014, GSSyT_r_*SSGT: β = 9.32, P = 0.0077). However, we did not detect any significant interaction (Supplementary Fig 8).

### Intervention outcome prediction using machine learning test

Lastly, we investigated whether a potential machine learning (ML) model could predict the response by integrating detailed clinical and genetic information from the individuals in the KONTAKT® trial. Participants were separated into two classes based on if they had responses to the treatment (n = 88 (47.3%)) or not (n = 98 (52.7%)) measured by parent-reported SRS at post-intervention. After evaluation, we applied linear support vector machine (Supplementary Information) on selected twelve features, including individual comorbidities, SSGT treatment, SRS and Clinical Global Impression (CGI) score, rare exome variants (number of rare damaging/developmental disorder-related variants), rare CNVs (number of genes in rare CNVs, rare CNV size) and GSSyT_r_ (Supplementary Fig 9). The average area under the receiver operating characteristic curve (AUC) and accuracy were 0.632 and 0.634 (Fig 4). The ML model was able to correctly predict individuals’ responses to interventions with an average of 60% sensitivity and 66.4% specificity (Fig 4).

**Figure 4.**
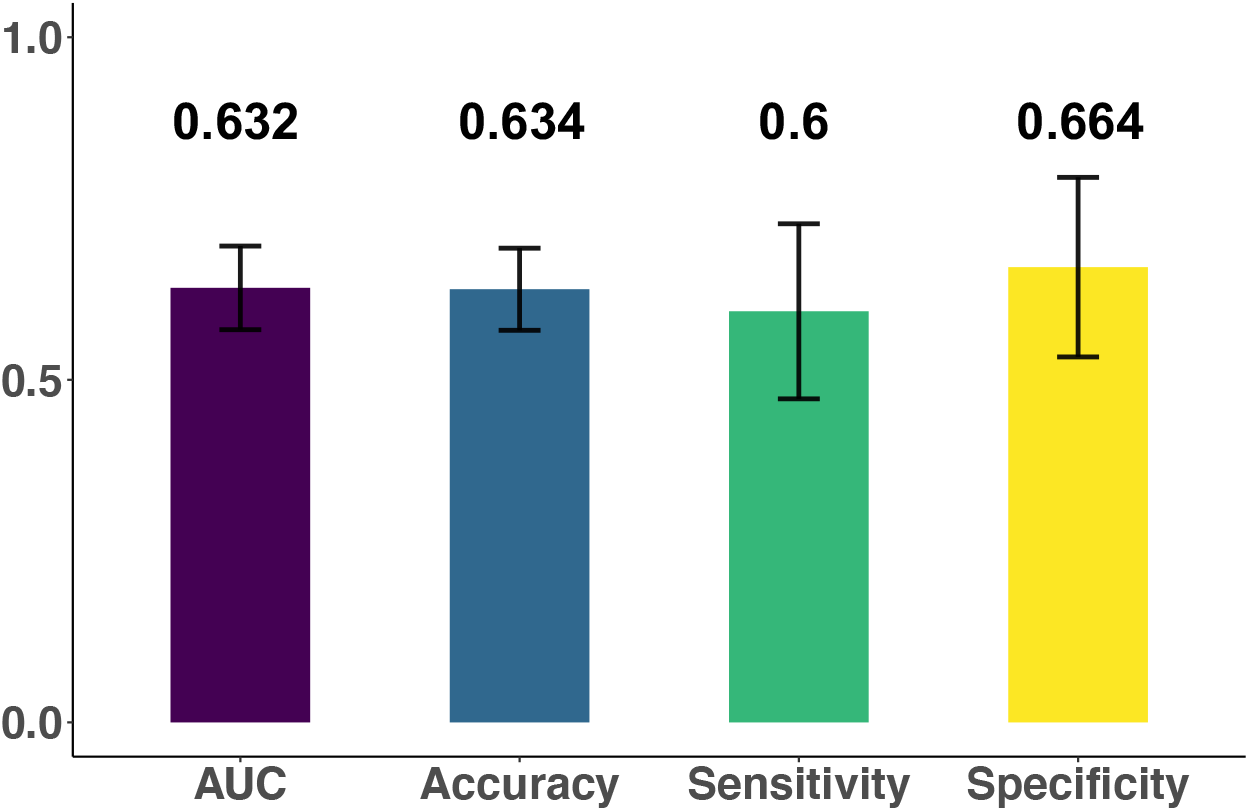
Performance of machine learning model measured by area under the receiver operating characteristic curve (AUC), accuracy, sensitivity and specificity

## Discussion

Most of the current interventions yield variable responses among autistic individuals. There is minimal knowledge of whether genetic information can be used to guide clinicians to individualize intervention decisions. A few case studies [26–28] and our earlier reports indicate a potential to identify an association between intervention outcome and genetic information [21,22]. Despite large-scale exome sequencing studies in ASD, no studies have directly connected ASD-related rare sequence variants to intervention outcomes. Here, we investigated whether carrier status of rare ASD-related sequence variants, categorized as variants of clinical significance (VCSs), variants of uncertain significance (VUSs), or variants in genes within synaptic transmission pathway, have an effect on outcomes following SSGT and standard care from a recent clinical trial. After variant prioritization and the association test, we found that individuals with VCS or VUS together showed limited social behavioral change across the entire cohort and further analysis pinpointed that this effect was especially evident for the standard care group. Furthermore, we showed that the effect was driven by changes in the social cognition subscale of the SRS. Additionally, the limiting effect of VCS was stronger than VUS in both the entire cohort and the standard care. As SSGT was an add-on intervention to the standard care in the KONTAKT® trial, the non-significant difference between carriers and non-carriers in the SSGT group might indicate a potentially beneficial effect of SSGT for carriers compared to standard care alone, which should be explored further. Earlier, we demonstrated that individuals with clinically significant CNVs had better outcomes in standard care and less improvements after SSGT [21]. Interestingly, in our pilot-scheme of genetic scores based on variant effects on genes within synaptic transmission pathway (GSSyT), we similarly showed that rare variant GSSyT (GSSyT_r_) and common variant GSSyT (GSSyT_c_) were associated with differential outcomes in standard care and SSGT groups. Further our ML model confirmed the importance of rare variants for outcome prediction and provided a feasible pattern for investigating the role of both clinical and genetic features on ASD-related treatments.

We also provided evidence that intellectually able autistic individuals carried genetic variants of clinical significance, as we detected that a minority of 4.4% individuals had VCS, which is within the range of other clinical exome sequencing studies [29–31]. Although the proportion is slightly lower than in earlier reports, it is important to note that these studies also included individuals with co-occurring intellectual disability, which is associated with an increased probability of VCS findings [29–31]. Combining clinically significant CNVs with the exome sequencing findings, the overall proportion of identifiable genetic etiology of autistic individuals in this cohort was 12.6%, demonstrating that this group of autistic individuals should also have access to clinical genetic testing. As no overlap was found between VCS and clinically significant CNV carriers, our results confirmed the complementary role of exome sequencing and chromosome microarray increasing the molecular diagnostic yield for individuals with ASD.

We demonstrated nine VCSs from different autosomal-dominant genes implicated in ASD and other NDDs. For example, variants from *CUL3* (OMIM* 603136) have been observed in individuals with pseudohypoaldosteronism (MIM* 614496) and contribute to ASD, social deficits, and anxiety-like behaviors [32–34]. Likewise, we found both clinically significant CNV and exome VCS affected *ASH1L* (OMIM* 607999) in our samples related to intellectual disability (MIM*617796), ASD, and Tourette syndrome [30,35]. This gene encodes a histone methyltransferase that associates with the transcribed region such as *Hox* genes, catalyzes H3K36 methylation, and plays important roles in neurodevelopment [36].

When assessing the distribution of rare and common variants in this cohort, we found that carriers of pathogenic variants and non-carriers had identical levels of PRS for ASD, which is in line with a recent study using a much larger cohort showing that the contribution from common genetic variants is similarly relevant to autistic individuals with or without a strongly acting de novo mutation [8]. In addition, Niemi *et al*. compared PRS between children with severe NDDs (e.g., global developmental delay, intellectual disability, cognitive impairment or learning disabilities, and ASD) with and without pathogenic variants and found no significant differences for any tested PRSs [37]. All these results suggest PRS and rare variants associated with ASD or other NDDs may contribute additively to the individual liability for the conditions. Similarly, we found no interaction effect of PRS for ASD and exome pathogenic variants on interventions. As a similar independent role for PRS and clinically significant CNV was shown earlier [22], our study indicated an additive or differential role for each type of variant contributing to the treatment outcomes.

There is a high genetic and clinical heterogeneity in ASD. Besides traditional association tests, an ML model can integrate measurements to recapitulate useful information to predict intervention outcomes. Using the KONTAKT® trial data, we built a prediction model to select the most contributing features to the intervention responses, including variables representing the complexity and severity of the disorder (comorbidity status, SRS and CGI scores), intervention conditions (SSGT intervention), and genetic liability especially rare variant related characteristics. These selected features from different domains could provide not only a prioritized list for future ASD investigations but more evidence for precise guidance into clinical practice of intervention decisions. When the linear model, especially the linear SVM, combined selected variables into outcomes, it outperformed the non-linear model showing 63.4% accuracy, illustrating the outcome was to a certain degree predictable instead of by chance (50%) to help clinicians decide whether an individual could have a response or not after adding SSGT as an add-on of standard care. Due to limited power, our study was inadequate to make predictions for each intervention subgroup. As no study has combined clinical and genetic factors to predict the outcomes of ASD interventions, the performance of prediction in our study could be considered a reference to compare after similar studies become available.

Limitations in our study should also be noticed. To begin with, although our study was based on the largest randomized controlled trial of SSGT in ASD to date, limited sample size restricted the power to identify intervention-associated genes and to stratify variant effect in subgroups. Exome sequencing studies of ASD have shown that both *de novo* and inherited rare variants are major components of genetic risk and are associated with cognitive phenotypes such as IQ [38]. However, due to the lack of parental information, our study could not distinguish *de novo* and inherited rare variants and seek out any differences between variant types. Furthermore, as the treatment methods are varied for ASD, we could not combine samples from other clinical cohorts and validate our findings. Although some completed and ongoing clinical trials testing the SSGT effect, exome sequencing data on the same cohorts are unavailable at present [39,40]. Therefore, without further validations, our results must be interpreted with caution. With increased genetic testing in clinics, converting genetic information into primary diagnosis and treatment, combined with clinical observation and biometric data, could proactively shape clinical decisions and long-term management [41]. Factors from different aspects should be considered when leveraging genetic data into practice, such as data interpretation, genetic counseling, ethical consideration, and economic constraints [41,42].

## Conclusions

In summary, we have demonstrated that exome sequencing can identify molecular diagnoses in an ASD cohort without intellectually disabled individuals, and genetic information reveals significant associations with intervention outcomes, although with variable effects in the SSGT and standard care groups. We also show that gene set based genetic scores could be further explored to determine meaningful subgroups and direct future studies on biological mechanisms. By developing an ML model, we confirm that individual outcomes could be predicted by clinical and genetic factors such as rare variants. Our study is a preliminary exploration of rare variants and a combination of different level variants on ASD intervention outcomes. Future clinical trials of different interventions in ASD should include genetic data collection to improve the use of molecular genetics beyond diagnoses for individualized intervention plans.

## Data Availability

All public data from databases and studies were described in the manuscript. The genetic data is accessible through the Swedish National Data Service (https://snd.gu.se/en) after necessary clearances.

## List of abbreviations

ASD: autism spectrum disorder
CNV: copy number variation
NDD: neurodevelopmental disorder
PRS: polygenic risk score
ML: machine learning
SSGT: social skills group training
ADHD: attention deficit hyperactivity disorder
SRS: Social Responsiveness Scale
VCS: variant of clinical significance
VUS: variant of uncertain significance
GSSyT: genetic score for synaptic transmission
GSSyT_c_: genetic score for synaptic transmission for common variant
GSSyT_r_: genetic score for synaptic transmission for rare variant
SVM: support vector machine
AUC: area under the receiver operating characteristic curve

## Declarations

### Ethics approval

Written informed consent was obtained from parents or legal guardians, and verbal consent from all participants. All the protocols and methods were followed in accordance with the Declaration of Helsinki and were approved by the ethical review board in Stockholm (Dnr 2012/385-31/4) and the clinical authorities of the two involved counties.

### Availability of data and materials

All public data from databases and studies were described in the manuscript. The genetic data is accessible through the Swedish National Data Service (https://snd.gu.se/en) after necessary clearances. Code related to variants prioritization and developmental disorder related variants selection is available at: https://github.com/DanyangLi107/vcf_varselect. Code related to machine learning test is available at: https://github.com/DanyangLi107/ml_kontakt. All statistical analyses were performed in R v3.6.3. Additional software and package information were illustrated in the Methods and Supplementary Information.

### Authors’ contributions

D.L and K.T conceived and planned the present study. D.L designed and performed the analyses with supervision from K.T. H.J and N.N provided overall suggestion and discussion for the analyses. M.B and A.A performed experiments of prioritized variants for Sanger validation. S.B and N.C-O conceived the original clinical trial. S.B is responsible for the clinical data and provided the expertise of the clinical measures together with N.C-O, and U.J. D.L and K.T wrote the paper with input from all authors. All authors approved the final version of the article.

## Acknowledgments

We thank the children, adolescents, and parents who participated in the study. Christina Coco, Oskar Flygare, Anders Görling, and Kerstin Andersson are acknowledged for their work in collecting the data and samples during the RCT. The authors are also thankful for the leads of child and adolescent psychiatry (Olav Bengtsson, Paula Liljeberg, Charlotta Wiberg Spangenberg, Peter Ericson, Karin Forler, Alkisti Nikolayidis Linderholm, all of Stockholm County Council), PRIMA Järva child and adolescent psychiatry (MaiBritt Giacobini), and child and adolescent habilitation services (Lars Kjellin, Moa Pellrud, of Örebro County Council) for organizational support. The DNA extraction was done at the KI Biobank. We thank Anna Gellerbring, Emma Sernstad, Emilia Ottosson and Valtteri Wirta from Clinical Genomics Stockholm core facility, Karolinska Institutet, and Science for Life Laboratory for exome sequencing experiments and variant calling process. The computation resources were provided by SNIC through Uppsala Multidisciplinary Center for Advanced Computational Science (UPPMAX). We would like to acknowledge the Swedish Bioinformatics Advisory Program at the National Bioinformatics Infrastructure Sweden, SciLifeLab, for bioinformatics consultation for data analysis.

## Reference

1. Finucane BM, Ledbetter DH, Vorstman JA. Diagnostic genetic testing for neurodevelopmental psychiatric disorders: closing the gap between recommendation and clinical implementation. Curr Opin Genet Dev. 2021;68:1–8.

2. Havdahl A, Niarchou M, Starnawska A, Uddin M, van der Merwe C, Warrier V. Genetic contributions to autism spectrum disorder. Psychol Med. Cambridge University Press; 2021;1–14. http://www.ncbi.nlm.nih.gov/pubmed/33634770

3. American Psychiatric Association. Diagnostic and Statistical Manual of Mental Disorders (DSM-5), Fifth Edition. Arlington, VA: American Psychiatric Publishing; 2013.

4. Miller DT, Adam MP, Aradhya S, Biesecker LG, Brothman AR, Carter NP, et al. Consensus Statement: Chromosomal Microarray Is a First-Tier Clinical Diagnostic Test for Individuals with Developmental Disabilities or Congenital Anomalies. Am J Hum Genet. 2010;86:749–64. https://www.sciencedirect.com/science/article/pii/S0002929710002089?via%3Dihub

5. Srivastava S, Love-Nichols JA, Dies KA, Ledbetter DH, Martin CL, Chung WK, et al. Meta-analysis and multidisciplinary consensus statement: exome sequencing is a first-tier clinical diagnostic test for individuals with neurodevelopmental disorders. Genet Med. 2019;21:2413–21. http://www.nature.com/articles/s41436-019-0554-6

6. Moreno-De-Luca D, Kavanaugh BC, Best CR, Sheinkopf SJ, Phornphutkul C, Morrow EM. Clinical genetic testing in autism spectrum disorder in a large community-based population sample. JAMA Psychiatry. 2020;77(9):979–81. https://jamanetwork.com/

7. Grove J, Ripke S, Als TD, Mattheisen M, Walters RK, Won H, et al. Identification of common genetic risk variants for autism spectrum disorder. Nat Genet. 2019;51:431–44. http://www.nature.com/articles/s41588-019-0344-8

8. Weiner DJ, Wigdor EM, Ripke S, Walters RK, Kosmicki JA, Grove J, et al. Polygenic transmission disequilibrium confirms that common and rare variation act additively to create risk for autism spectrum disorders. Nat Genet. 2017;49:978–85.

9. Johannessen J, Nærland T, Hope S, Torske T, Høyland AL, Strohmaier J, et al. Parents’ attitudes toward clinical genetic testing for autism spectrum disorder—data from a norwegian sample. Int J Mol Sci. 2017;18:1078. www.mdpi.com/journal/ijms

10. Berry-Kravis E, Des Portes V, Hagerman R, Jacquemont S, Charles P, Visootsak J, et al. Mavoglurant in fragile X syndrome: Results of two randomized, double-blind, placebo-controlled trials. Sci Transl Med. 2016;8:321ra5. www.ScienceTranslationalMedicine.org

11. Satterstrom FK, Kosmicki JA, Wang J, Breen MS, De Rubeis S, An JY, et al. Large-Scale Exome Sequencing Study Implicates Both Developmental and Functional Changes in the Neurobiology of Autism. Cell. 2020;180:568–584.e23.

12. Ruzzo EK, Pérez-Cano L, Jung J-Y, Wang L, Kashef-Haghighi D, Hartl C, et al. Inherited and De Novo Genetic Risk for Autism Impacts Shared Networks. Cell. 2019;178:850-866.e26. https://linkinghub.elsevier.com/retrieve/pii/S0092867419307809

13. De Rubeis S, He X, Goldberg AP, Poultney CS, Samocha K, Ercument Cicek A, et al. Synaptic, transcriptional and chromatin genes disrupted in autism. Nature. 2014;515:209–15. http://www.nature.com/articles/nature13772

14. Krishnan A, Zhang R, Yao V, Theesfeld CL, Wong AK, Tadych A, et al. Genome-wide prediction and functional characterization of the genetic basis of autism spectrum disorder. Nat Neurosci. 2016;19:1454–62.

15. Asif M, Martiniano HFMC, Marques AR, Santos JX, Vilela J, Rasga C, et al. Identification of biological mechanisms underlying a multidimensional ASD phenotype using machine learning. Transl Psychiatry. 2020;10:43. https://doi.org/10.1038/s41398-020-0721-1

16. Chen S, Fragoza R, Klei L, Liu Y, Wang J, Roeder K, et al. An interactome perturbation framework prioritizes damaging missense mutations for developmental disorders. Nat Genet. 2018;50:1032–40. https://doi.org/10.1038/s41588-018-0130-z

17. Lenhard F, Sauer S, Andersson E, Månsson KN, Mataix-Cols D, Rück C, et al. Prediction of outcome in internet-delivered cognitive behaviour therapy for paediatric obsessive-compulsive disorder: A machine learning approach. Int J Methods Psychiatr Res. 2018;27:e1576. http://doi.wiley.com/10.1002/mpr.1576

18. Kautzky A, Baldinger P, Souery D, Montgomery S, Mendlewicz J, Zohar J, et al. The combined effect of genetic polymorphisms and clinical parameters on treatment outcome in treatment-resistant depression. Eur Neuropsychopharmacol. 2015;25:441–53. http://dx.doi.org/10.1016/j.euroneuro.2015.01.001

19. Chancel R, Miot S, Dellapiazza F, Baghdadli A. Group-based educational interventions in adolescents and young adults with ASD without ID: a systematic review focusing on the transition to adulthood. Eur Child Adolesc Psychiatry. 2020; https://doi.org/10.1007/s00787-020-01609-1

20. Choque Olsson N, Flygare O, Coco C, Görling A, Råde A, Chen Q, et al. Social Skills Training for Children and Adolescents With Autism Spectrum Disorder: A Randomized Controlled Trial. J Am Acad Child Adolesc Psychiatry. 2017;56:585–92. https://linkinghub.elsevier.com/retrieve/pii/S0890856717302022

21. Tammimies K, Li D, Rabkina I, Stamouli S, Becker M, Nicolaou V, et al. Association between Copy Number Variation and Response to Social Skills Training in Autism Spectrum Disorder. Sci Rep. 2019;9:9810. http://www.nature.com/articles/s41598-019-46396-1

22. Li D, Choque-Olsson N, Jiao H, Norgren N, Jonsson U, Bölte S, et al. The influence of common polygenic risk and gene sets on social skills group training response in autism spectrum disorder. npj Genomic Med. 2020;5:45. http://dx.doi.org/10.1038/s41525-020-00152-x

23. Constantino JN, Gruber CP. Social Responsiveness Scale (SRS). Los Angeles: Western Psychological Services; 2005.

24. Stranneheim H, Engvall M, Naess K, Lesko N, Larsson P, Dahlberg M, et al. Rapid pulsed whole genome sequencing for comprehensive acute diagnostics of inborn errors of metabolism. BMC Genomics. 2014;15:1090. http://bmcgenomics.biomedcentral.com/articles/10.1186/1471-2164-15-1090

25. Richards S, Aziz N, Bale S, Bick D, Das S, Gastier-Foster J, et al. Standards and guidelines for the interpretation of sequence variants: a joint consensus recommendation of the American College of Medical Genetics and Genomics and the Association for Molecular Pathology. Genet Med. 2015;17:405–23. http://www.nature.com/articles/gim201530

26. Cucinotta F, Ricciardello A, Turriziani L, Calabrese G, Briguglio M, Boncoddo M, et al. FARP-1 deletion is associated with lack of response to autism treatment by early start denver model in a multiplex family. Mol Genet Genomic Med. 2020;8:e1373. https://onlinelibrary.wiley.com/doi/10.1002/mgg3.1373

27. Serret S, Thümmler S, Dor E, Vesperini S, Santos A, Askenazy F. Lithium as a rescue therapy for regression and catatonia features in two SHANK3 patients with autism spectrum disorder: Case reports. BMC Psychiatry. 2015;15:107. http://bmcpsychiatry.biomedcentral.com/articles/10.1186/s12888-015-0490-1

28. Pini G, Scusa MF, Benincasa A, Bottiglioni I, Congiu L, Vadhatpour C, et al. Repeated Insulin-Like Growth Factor 1 Treatment in a Patient with Rett Syndrome: A Single Case Study. Front Pediatr. 2014;2:52. http://journal.frontiersin.org/article/10.3389/fped.2014.00052/abstract

29. Guo H, Duyzend MH, Coe BP, Baker C, Hoekzema K, Gerdts J, et al. Genome sequencing identifies multiple deleterious variants in autism patients with more severe phenotypes. Genet Med. 2019;21:1611–20.

30. Tammimies K, Marshall CR, Walker S, Kaur G, Thiruvahindrapuram B, Lionel AC, et al. Molecular diagnostic yield of chromosomal microarray analysis and whole-exome sequencing in children with autism spectrum disorder. JAMA. 2015;314:895–903.

31. Iossifov I, O’roak BJ, Sanders SJ, Ronemus M, Krumm N, Levy D, et al. The contribution of de novo coding mutations to autism spectrum disorder. Nature. 2014;515:216–221.

32. Dong Z, Chen W, Chen C, Wang H, Cui W, Tan Z, et al. CUL3 Deficiency Causes Social Deficits and Anxiety-like Behaviors by Impairing Excitation-Inhibition Balance through the Promotion of Cap-Dependent Translation. Neuron. 2020;105:475–490.e6. https://doi.org/10.1016/j.neuron.2019.10.035

33. O‘Roak BJ, Vives L, Girirajan S, Karakoc E, Krumm N, Coe BP, et al. Sporadic autism exomes reveal a highly interconnected protein network of de novo mutations. Nature. 2012;485:246–50. https://www.nature.com/articles/nature10989

34. Boyden LM, Choi M, Choate KA, Nelson-Williams CJ, Farhi A, Toka HR, et al. Mutations in kelch-like 3 and cullin 3 cause hypertension and electrolyte abnormalities. Nature. 2012;482:98–102. https://www.nature.com/articles/nature10814

35. Liu S, Tian M, He F, Li J, Xie H, Liu W, et al. Mutations in ASH1L confer susceptibility to Tourette syndrome. Mol Psychiatry. 2020;25:476–90. https://doi.org/10.1038/s41380-019-0560-8

36. Okamoto N, Miya F, Tsunoda T, Kato M, Saitoh S, Yamasaki M, et al. Novel MCA/ID syndrome with ASH1L mutation. Am J Med Genet Part A. 2017;173:1644–8. http://doi.wiley.com/10.1002/ajmg.a.38193

37. Niemi MEK, Martin HC, Rice DL, Gallone G, Gordon S, Kelemen M, et al. Common genetic variants contribute to risk of rare severe neurodevelopmental disorders. Nature. 2018;562:268–271. https://doi.org/10.1038/s41586-018-0566-4

38. Iakoucheva LM, Muotri AR, Sebat J. Getting to the Cores of Autism. Cell. 2019;178:1287–98. https://www.sciencedirect.com/science/article/pii/S0092867419308360

39. Freitag CM, Jensen K, Elsuni L, Sachse M, Herpertz-Dahlmann B, Schulte-Rüther M, et al. Group-based cognitive behavioural psychotherapy for children and adolescents with ASD: the randomized, multicentre, controlled SOSTA - net trial. J Child Psychol Psychiatry. 2016;57:596–605. http://doi.wiley.com/10.1111/jcpp.12509

40. Gates JA, Kang E, Lerner MD. Efficacy of group social skills interventions for youth with autism spectrum disorder: A systematic review and meta-analysis. Clin Psychol Rev. 2017;52:164–81. https://www.sciencedirect.com/science/article/pii/S027273581630352X?via%3Dihub

41. Katsanis SH, Katsanis N. Molecular genetic testing and the future of clinical genomics. Nat Rev Genet. 2013;14:415–26. http://www.nature.com/articles/nrg3493

42. Horton RH, Lucassen AM. Recent developments in genetic/genomic medicine. Clin. Sci. 2019;133:697–708. https://doi.org/10.1042/CS20180436

