## Supplementary Information for "Rare variants in the outcome of social skills group training for autism"

### Supplementary Methods

#### Participants

The inclusion and exclusion criteria, participant characteristics and the trial outcome have been reported earlier [1]. Demographic and clinical information was available from the social skills group training (SSGT) KONTAKT trial including individual characteristics (IQ, age, sex), comorbidity diagnoses (attention deficit hyperactivity disorder (ADHD), depression, anxiety, others), treatments (SSGT, or standard care in the following subgroups: cognitive behavior therapy, pharmaceutical treatment, individual counseling) and autism-related scales (Social Responsiveness Scale (SRS), Autism Diagnostic Observation Schedule (ADOS) social communication total scores, Developmental Disabilities Modification of Children's Global Assessment Scale (DD-CGAS), Clinical Global Impression (CGI), and Adaptive Behavior Assessment System (ABAS)) [1].

#### Exome sequencing

Saliva-derived genomic DNA (50 ng) of each sample was used for library preparation with the Twist Human Core Exome v1.3 Enrichment Kit (Twist Bioscience). The following modifications: xGen Dual Index UMI adapters (6-nt unique molecular identifiers (UMI), 0.6 mM, Integrated DNA Technologies) were used for the ligation and xGen Library Amp Primer (2 mM, Integrated DNA Technologies) was used for PCR amplification (10 cycles). Target enrichment was performed in a multiplex fashion with a library amount of 187.5 ng (8-plex). The libraries were hybridized to Exome probes v1.3 (Twist Bioscience), xGen Universal Blockers - TS Mix (Integrated DNA Technologies) and COT Human DNA (Life Technologies) for 16-19 hours. The post-capture PCR was performed with xGen Library Amp Primer (0.5 mM, Integrated DNA Technologies) for eight cycles. Quality control was performed with the Qubit dsDNA HS assay (Invitrogen) and TapeStation HS D1000 assay (Agilent). Sequencing was done on NovaSeq 6000 using paired-end 2 × 150 readout. Demultiplexing was done using Casava v2.20. Exome sequencing was performed at the Clinical Genomics Stockholm core facility, Karolinska Institutet, and Science for Life Laboratory, Stockholm, Sweden.

#### Developmental disorder-related genes curation

As there is no consensus on what genes are clinically evaluated for individuals with ASD [2], we generalized a developmental disorder-related gene list from different resources. We used three online databases: Simons Foundation Autism Research Initiative (SFARI, https://gene.sfari.org/), DatabasE of genomiC varIation and Phenotype in Humans using Ensembl Resources (DECIPHER, https://decipher.sanger.ac.uk/ddd#overview), and Human Phenotype Ontology (HPO, https://hpo.jax.org/app/) to obtain genes linked to the etiology of ASD and developmental disorders (DDs). Genes from DECIPHER, SFARI category 1, 2, and syndromic, as well as behavioral abnormality (HP:0000708) and cognitive impairment (HP:0100543) in HPO were selected. Additionally, genes significantly associated with ASD, intellectual disability, and developmental delay were extracted from five recent genome/ exome sequencing and chromosome microarray studies [3–7]. Online Mendelian Inheritance in Man (OMIM, https://www.omim.org/) database and Clinical Genomics Database (CGD, https://research.nhgri.nih.gov/CGD/) were used for curating the inheritance mode of the gene. Gene scores, including pLI and missense-Z were matched from genome aggregation database (gnomAD, https://gnomad.broadinstitute.org/). All data were downloaded in June 2019, and the final selected DD genes were listed in Supplementary Table 1.

**Variant prioritization**

Quality control and rare variant selection

The called variants were first filtered based on quality and minor allele frequency. Variants were defined as low quality if: i) Variant Quality Score Recalibration was not PASS, ii) read depth < 10, iii) QualByDepth < 2, iv) RMSMappingQuality < 40, v) loss-of-function (LOF) variants were not called by at least two callers (SAMtools, GATK, freebayes). We further categorized variants as common based on their minor allele frequency > 0.1% in any of the four data resources: 1000 Genomes, SweGen [8], gnomAD and ExAC [9], as well as the minor allele frequency > 1% in our in-house data. We proceeded with the following analysis using only rare variants that passed the quality filters.

Variant effect and clinical significance

Variant effect, defined by the Sequence Ontology (http://www.sequenceontology.org/), was annotated by Ensembl Variant Effect Predictor (VEP) v92. For variants annotated as missense, we further defined them as damaging missense if they followed at least 4 of these 6 criteria: i) SIFT [10] = 'deleterious' or 'deleterious low confidence', ii) PolyPhen2 [11] = 'possibly damaging' or 'probably damaging', iii) MPC [12] ≥ 2.0, iv) CADD [13] ≥ 20.0, v) |SPIDEX| [14] ≥ 2.0, vi) phyloP 100 vertebrates [15] ≥ 2.0. After quality control, qualified rare variants with the effect of LOF (splice donor/acceptor, frameshift, start lost, stop gained) or damaging missense were prioritized as rare deleterious variants for clinical significance assessment.

Rare deleterious variants that were located in developmental disorder-related genes and in accordance with the gene inheritance pattern were considered developmental disorder-related rare variants. To assess clinically significant variants, we further prioritized these developmental disorder -related rare variants using the following conditions: for LOF variants, i) genes where variants located had pLI score [16] > 0.9, ii) variants were not clustered (within a distance of 5bp) and were not in repeat regions if they were indels; for missense variants, i) genes where variants located had missense-Z score [16] > 3.09, ii) variants were recorded as either MPC [12] ≥ 2.0 or |SPIDEX| [14] ≥ 2. Furthermore, a detailed evaluation to categorize the variants to pathogenic/likely pathogenic, uncertain clinical effect, or benign/likely benign based on ACMG guideline was done [17], including variants position confirmation, clinical effect evaluation of variant located genes, and variant effect reported from other clinical studies. Both pathogenic and likely pathogenic variants were classified as variants of clinical significance (VCSs). All VCSs and variants of uncertain significance (VUSs) were validated by Sanger sequencing. Parental DNA was not available to investigate the inheritance of the variants.

**Genetic score for synaptic transmission (GSSyT)**

To test a pilot scheme for analyzing gene set specific genetic load, we constructed genetic scores for synaptic transmission (SyT) genes (GSSyT). Our earlier study based on common variants had shown nominal association with different synaptic gene sets and thus synaptic transmission genes were chosen for our pilot scheme [18]. The synaptic transmission gene set was acquired from Gene Ontology (GO:0007268) including 727 protein-coding genes. For the background set, we used all brain-expressed genes obtained from The human brain – The Human Protein Atlas (HPA, https://www.proteinatlas.org/humanproteome/brain) which included 15,157 protein-coding brain genes selected by the HPA with regional classification based on FAMTOM5 and GTEx. All data were downloaded in April 2020.

To calculate GSSyT based on the rare genetic variants ($GSSyT_r$), we utilized both rare deleterious exome variants and rare CNVs found in the individuals. Rare CNVs spanning at least 25 kb were called from CytoScan HD microarray previously [19]. As the sizes of CNVs varied, we separately summed the number of synaptic transmission genes and brain-expressed genes that the rare CNVs overlapped with, in each individual. For rare deleterious exome variants, we first calculated the number of variants in synaptic transmission genes and the number of affected synaptic transmission genes in each individual and found they were highly correlated (Pearson correlation, r = 1.00). A high correlation was also shown in the number of rare variants in brain-expressed genes, and the number of brain-expressed genes affected by these variants (Pearson correlation, r = 0.998). Therefore, we used the number of genes having rare deleterious exome variants in the two gene sets to combine with rare CNV genes. As no individual had synaptic genes affected by both rare CNVs and exome variants, and only two individuals each had one brain gene affected by the two types of rare variants, the number of synaptic genes captured by two kinds of rare variants were added and divided by the number of brain-expressed genes with rare variants (Equation 1). The ratio between the affected synaptic transmission genes to the affected brain-expressed genes was standardized to have mean = 0 and standard deviation = 1 for statistical analysis.

$$\mathrm{GSSyT_r} = \frac{\mathrm{SyT_{rc}} + \mathrm{SyT_{re}}}{\mathrm{Brain_{rc}} + \mathrm{Brain_{re}}} \quad (1)$$

*- Where $SyT_{rc}$ and $SyT_{re}$ is the number of synaptic transmission (SyT) genes affected by rare CNVs and rare deleterious exome variants. $Brain_{rc}$ and $Brain_{re}$ is the number of brain-expressed genes affected by rare CNVs and rare deleterious exome variants.*

Next, we generated GSSyT for the common variants ($GSSyT_c$). We acquired common variant data from the previous study including 5,126,694 SNPs after imputation [18]. The calculation of $GSSyT_c$ was based on set based polygenic risk score (PRS) using a new feature of PRSice named PRSet (https://www.prsice.info/quick_start_prset/). We chose the largest ASD GWAS of Grove et al. [20] as reference data downloaded from Psychiatric Genomics Consortium (https://www.med.unc.edu/pgc/download-results/asd/) to calculate set based PRS for ASD using multiple P-thresholds (Pts, $10^{-5}$, $10^{-4}$, $10^{-3}$, 0.01, 0.05, 0.10, 0.50, 1.00). Common variants of each Pt were allocated to synaptic transmission and brain-expressed gene sets to obtain PRSs separately. The ratio between synaptic transmission and brain PRSs was represented as $GSSyT_c$ (Equation 2) and further standardized (mean = 0 and standard deviation = 1) for association test.

$$\mathrm{GSSyT_c} = \frac{\mathrm{SyT_{PRS}}}{\mathrm{Brain_{PRS}}} \quad (2)$$

**Statistical analyses**

All statistical analyses were performed in R v3.6.3. We used one-way ANOVA to identify any statistically significant differences in IQ, pre-intervention SRS and ADOS-G in carriers of VCS/VUS and non-carriers. We also used additional genetic information including clinically significant CNVs and PRS for ASD (Pt 0.5) from our earlier studies [18,19]. The distribution of carriers with CS CNVs or exome VCSs/VUSs in the cohort was tested by Pearson's $\chi^2$ test. The distribution of ASD PRS was assessed between individuals with or without VCSs/VUSs using one-way ANOVA. The correlation of $GSSyT_c$ and $GSSyT_r$ in the synaptic transmission gene set was analyzed by Pearson correlation. The difference of $GSSyT_c$ and $GSSyT_r$ in the two treatment groups were tested using Student t-test.

Association analysis between VCS and intervention outcomes

We applied mixed linear model to assess whether exome VCS/VUS carrier status was associated with intervention outcome measured by parent-reported SRS total score and its four subscales (social awareness, social cognition, social motivation, social communication). Since we have shown earlier that age group, sex, clinically significant CNVs, and PRS for ADHD (Pt 1.0) affect intervention outcomes [1,18,19], and PRS for ADHD (Pt 1.0) and ASD (Pt 0.5) also have a high correlation [18], we included a three-way interaction: carrier status*time points*interventions together with all lower order interactions, age, sex, PRS for ADHD (Pt 1.0), clinically significant CNVs as fixed factors, and the clinics in which the individuals had participated in the intervention, and individual IDs as random factors in the model. The three-way interaction represented the role of exome VCS/VUS on SRS measured outcome in the SSGT group compared with the outcome in the standard care group. The two-way interaction: carrier status*time points showed the effect of exome VCS/VUS on the outcomes of both intervention groups at post-intervention and follow-up. In addition, to measure the separate effect of exome VCS/VUS on SSGT or standard care, the two-way interaction of carrier status*time points was tested using the same mixed linear model in SSGT and standard care subgroups separately. Furthermore, we also performed linear regression as a secondary model to test whether exome VCS/VUS and ASD PRS (Pt 0.5) independently affect the intervention outcomes. The changes of SRS at post-intervention and follow-up compared to pre-intervention were used as outcomes. We added the three-way interaction: exome variant*PRS*interventions, and adjusted sex and age in the model. Least-square means, coefficient values, and their 95% confidence interval (CI) were reported from the models.

GSSyT associations

Similarly, we conducted mixed linear model to investigate the effect of the variants from SyT on the intervention outcomes. We combined two three-way interactions of $GSSyT_r$*time points*interventions and $GSSyT_c$*time points*interventions, together with age, sex as fixed factors, and same random factors as earlier. The SRS total score reported by the parents was again used as the main outcome. At first, we calculated the explained variance for common variants at different Pts performed by "MuMIn" package v1.43.17. Only the set based PRS with the most explained Pt (Pt 0.01) was selected in the mixed linear model (marginal $R^2$ = 0.016). We also tested the effect of $GSSyT_c$ and $GSSyT_r$ in two intervention subgroups using the two-way interaction of GSSyT*time points. Furthermore, linear regression was performed to test the interaction of $GSSyT_c$ and $GSSyT_r$ using the change in SRS total score as well as sex and age as cofactors. Coefficient values and 95% CI were estimated in the models.

**Machine learning prediction**

<u>Outcome and features</u>

As an outcome for our machine learning prediction, a binary outcome for clinical improvement was calculated. The prediction model was constructed using the following participant characteristics: IQ, age, sex, comorbidity diagnoses, treatments, and autism-related scales assessed at pre-treatment. Furthermore, the following genetic information was used: information based on rare CNVs (CNV size, carrier status of clinically significant and large size (> 500 kb) CNVs, number of genes in CNVs), information from rare single nucleotide variations (SNVs)/indels (carrier status of exome VCS/VUS, number of rare damaging SNVs/indels and developmental disorder-related rare SNVs/indels, $GSSyT_r$), and information based on common variants (PRS for ADHD (Pt 1.0), PRS for ASD (Pt 0.5), $GSSyT_c$ (Pt 0.01)). Detailed feature information is listed in Supplementary Table 2. Missing values in each feature were imputed using k-Nearest Neighbor (k = 5). All features were then standardized (mean = 0, standard deviation = 1) to eliminate the scale difference.

<u>Model strategy</u>

We performed nested cross-validation to avoid model overfitting and information leakage by splitting total data into training and validation sets using outer cycle (10-fold) and then splitting the training set again into training and test data using inner cycle (5-fold). Linear support vector machine (SVM) was employed for prediction. At first, we implemented recursive feature elimination to rank most important features. This method aims to select features by recursively considering smaller and smaller sets of features given an external estimator that assigns weights to features such as the coefficients. We used inner cycle cross-validation to select features which were ranked first and then calculated selected frequency of each feature after outer cycle cross-validation finished. Then we categorized features into different sets based on their selected frequency. Different feature sets from the most to the least selected were sequential added in the model to repeat hyperparameters C using grid search with different values (C: [0.001, 0.01, 0.1, 1, 10]). After testing all the combinations, the hyperparameter value and the included feature sets with the highest model performance from the inner cycle cross-validation were applied to the outer cycle. Each class weight was balanced to adjust the weight inversely proportional to outcome frequency from the data. Final performance metrics were averaged across outer cycle.

Performance measures

We used area under the receiver operating characteristic curve (AUC) as model performance measurement to select hyperparameters and features in the inner cycle cross-validation. Similarly, during both inner and outer cross-validation, model with the best AUC in the training dataset was used to predict outcomes in the test and validation set. Other metrics, such as sensitivity, specificity, and accuracy were evaluated at each outer cycle cross-validation.

Alternative models

In addition to linear SVM, logistic regression and non-linear SVM (radial basis functions (RBF) kernel) were tested for outcome prediction applying same nested cross-validation and model measures. For feature selection, we calculated average coefficient of each feature in logistic regression and implemented sequential forward floating selection in RBF SVM. Both models used grid search to determine the best hyperparameters. However, after hyperparameter tuning and feature selection, the performance of both models performed worse compared to linear SVM described above (logistic regression: average $AUC_{max}$ = 0.622, RBF SVM: average $AUC_{max}$ = 0.604). In addition, the performance was declined using the change in SRS score (either responded or not) from pre-treament to follow-up (best performance linear SVM $AUC_{max}$ = 0.579). Therefore, in this paper, we only show detailed prediction results of linear SVM at post-intervention.

**References**

1. Choque Olsson N, Flygare O, Coco C, Görling A, Råde A, Chen Q, et al. Social Skills Training for Children and Adolescents With Autism Spectrum Disorder: A Randomized Controlled Trial. J Am Acad Child Adolesc Psychiatry. 2017;56:585–92. https://linkinghub.elsevier.com/retrieve/pii/S0890856717302022

2. Hoang N, Buchanan JA, Scherer SW. Heterogeneity in clinical sequencing tests marketed for autism spectrum disorders. npj Genomic Med. 2018;3:27. www.nature.com/npjgenmed

3. Sanders SJ, He X, Willsey AJ, Ercan-Sencicek AG, Samocha KE, Cicek AE, et al. Insights into Autism Spectrum Disorder Genomic Architecture and Biology from 71 Risk Loci. Neuron. 2015;87:1215–33. http://dx.doi.org/10.1016/j.neuron.2015.09.016

4. C Yuen RK, Merico D, Bookman M, L Howe J, Thiruvahindrapuram B, Patel R V, et al. Whole genome sequencing resource identifies 18 new candidate genes for autism spectrum disorder. Nat Neurosci. 2017;20:602–11. http://www.nature.com/articles/nn.4524

5. Satterstrom FK, Kosmicki JA, Wang J, Breen MS, De Rubeis S, An JY, et al. Large-Scale Exome Sequencing Study Implicates Both Developmental and Functional Changes in the Neurobiology of Autism. Cell. 2020;180:568-84.e23.
6. Coe BP, Stessman HAF, Sulovari A, Geisheker MR, Bakken TE, Lake AM, et al. Neurodevelopmental disease genes implicated by de novo mutation and copy number variation morbidity. Nat Genet. 2019;51:106–16. http://www.nature.com/articles/s41588-018-0288-4
7. Pinto D, Delaby E, Merico D, Barbosa M, Merikangas A, Klei L, et al. Convergence of Genes and Cellular Pathways Dysregulated in Autism Spectrum Disorders. Am J Hum Genet. 2014;94:677–94. https://www.sciencedirect.com/science/article/pii/S0002929714001505?via%3Dihub#app2
8. Ameur A, Dahlberg J, Olason P, Vezzi F, Karlsson R, Martin M, et al. SweGen: A whole-genome data resource of genetic variability in a cross-section of the Swedish population. Eur J Hum Genet. 2017;25:1253–60. https://swefreq.nbis.se.
9. Karczewski KJ, Francioli LC, Tiao G, Cummings BB, Alföldi J, Wang Q, et al. The mutational constraint spectrum quantified from variation in 141,456 humans. Nature. Nature Research; 2020;581:434–43. https://doi.org/10.1038/s41586-020-2308-7
10. Ng Pauline C, Steven H. Predicting Deleterious Amino Acid Substitutions. Genome Res. 2001;11:863–74. http://genome.cshlp.org/content/11/5/863.abstract
11. Adzhubei IA, Schmidt S, Peshkin L, Ramensky VE, Gerasimova A, Bork P, et al. A method and server for predicting damaging missense mutations. Nat Methods. 2010;7:248–9. http://dx.doi.org/10.1038/nmeth0410-248
12. Samocha KE, Kosmicki JA, Karczewski KJ, O'Donnell-Luria AH, Pierce-Hoffman E, MacArthur DG, et al. Regional missense constraint improves variant deleteriousness prediction. bioRxiv. 2017;148353. https://www.biorxiv.org/content/early/2017/06/12/148353
13. Kircher M, Witten DM, Jain P, O'roak BJ, Cooper GM, Shendure J. A general framework for estimating the relative pathogenicity of human genetic variants. Nat Genet. 2014;46:310–5. https://www.nature.com/articles/ng.2892
14. Xiong HY, Alipanahi B, Lee LJ, Bretschneider H, Merico D, Yuen RKC, et al. The human splicing code reveals new insights into the genetic determinants of disease. Science. 2015;347:1254806.
15. Pollard KS, Hubisz MJ, Rosenbloom KR, Siepel A. Detection of nonneutral substitution rates on mammalian phylogenies. Genome Res. 2010;20:110–21. http://www.ncbi.nlm.nih.gov/pubmed/19858363
16. Lek M, Karczewski KJ, Minikel E V., Samocha KE, Banks E, Fennell T, et al. Analysis of protein-coding genetic variation in 60,706 humans. Nature. 2016;536:285–91.
17. Richards S, Aziz N, Bale S, Bick D, Das S, Gastier-Foster J, et al. Standards and guidelines for the interpretation of sequence variants: a joint consensus recommendation of the American College of Medical Genetics and Genomics and the Association for Molecular Pathology. Genet Med. 2015;17:405–23. http://www.nature.com/articles/gim201530
18. Li D, Choque-Olsson N, Jiao H, Norgren N, Jonsson U, Bölte S, et al. The influence of common polygenic risk and gene sets on social skills group training response in autism spectrum disorder. npj Genomic Med. 2020;5:45. http://dx.doi.org/10.1038/s41525-020-00152-x
19. Tammimies K, Li D, Rabkina I, Stamouli S, Becker M, Nicolaou V, et al. Association between Copy Number Variation and Response to Social Skills Training in Autism Spectrum Disorder. Sci Rep. 2019;9:9810. http://www.nature.com/articles/s41598-019-46396-1
20. Grove J, Ripke S, Als TD, Mattheisen M, Walters RK, Won H, et al. Identification of common genetic risk variants for autism spectrum disorder. Nat Genet. 2019;51:431–44. http://www.nature.com/articles/s41588-019-0344-8

**Supplementary Figures**

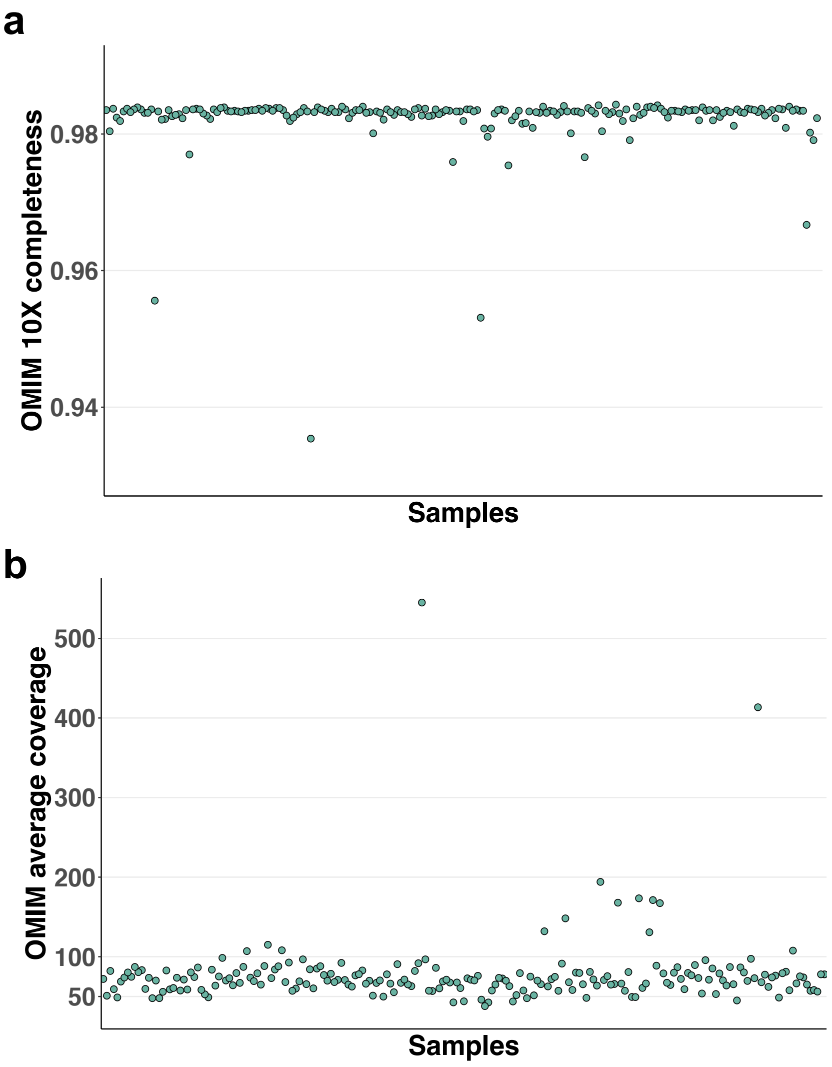

**Supplementary Figure 1**. Quality of exome sequencing in each sample. **a**. OMIM gene average coverage. **b**. OMIM gene 10X completeness.

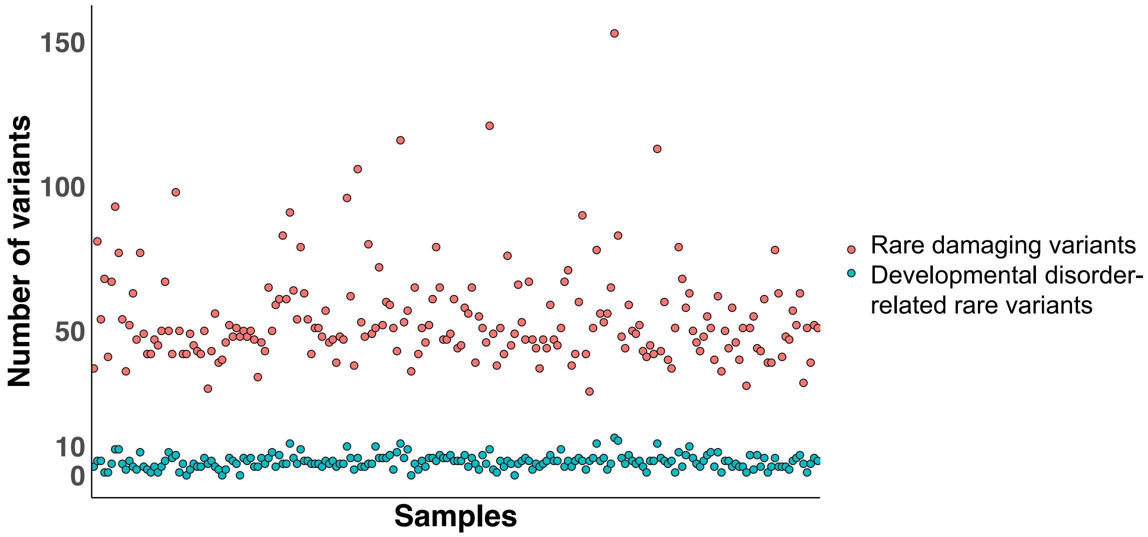

**Supplementary Figure 2**. The number of rare damaging exome variants and developmental disorder-related rare exome variants in each sample.

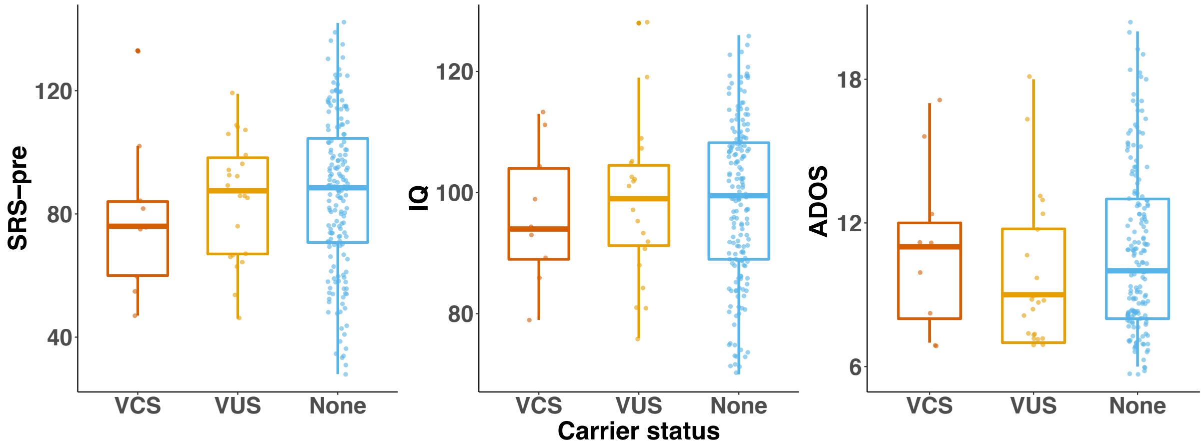

**Supplementary Figure 3**. Distribution of carrier status in Social Responsiveness Scale at pre-intervention (SRS-pre), IQ level and Autism Diagnostic Observation Schedule (ADOS) score. VUS: variant of uncertain significance, VCS: variant of clinical significance

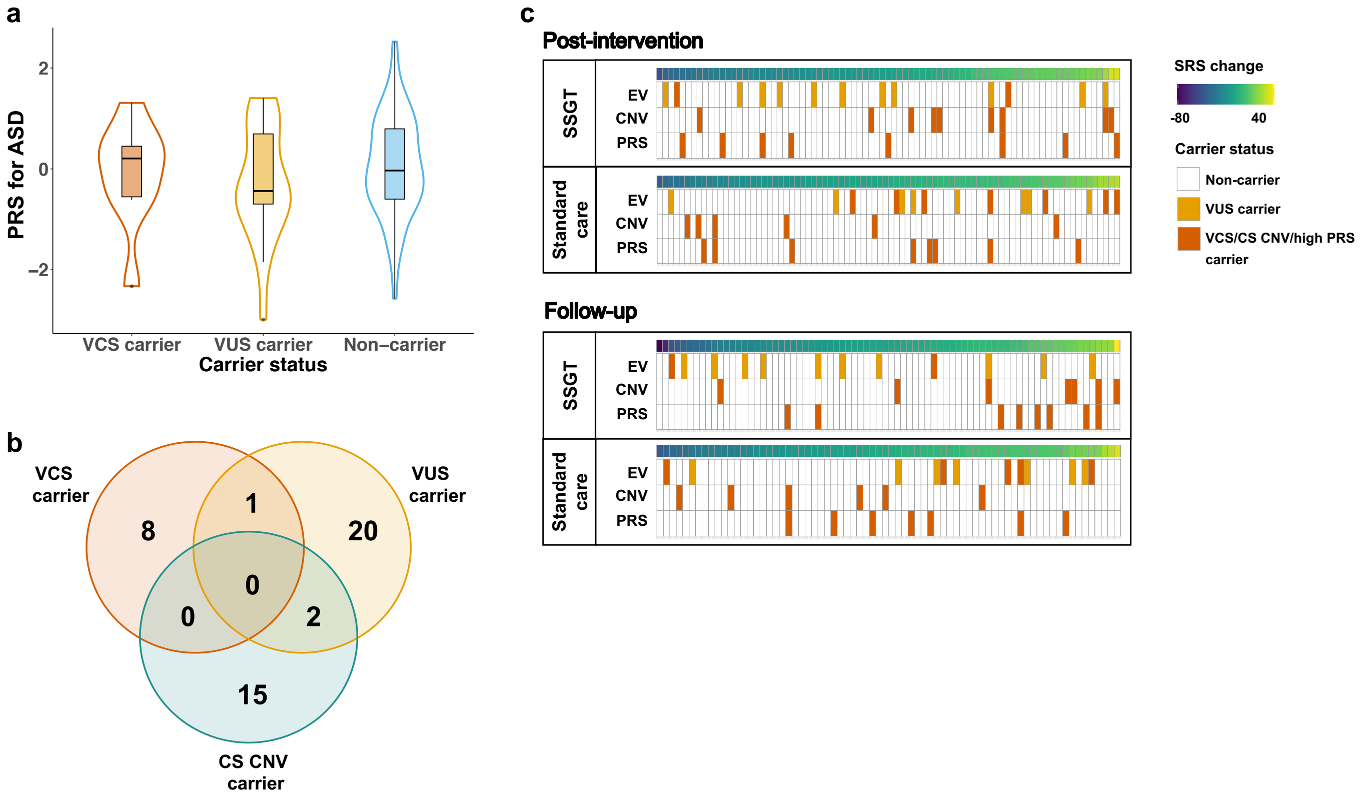

**Supplementary Figure 4**. Distribution of the prioritized genetic variants within the study cohort. **a**. Distribution of Autism Spectrum Disorder (ASD) polygenic risk score (PRS) (P-threshold 0.5) in VCS/VUS carriers and non-carriers **b**. Number and overlap of carriers with VCS/VUS and clinically significant (CS) copy number variant (CNV). **c**. Distribution of CS CNV, exome VCS/VUS, and 10% highest PRS for ASD (P-threshold 0.5) carriers. Individuals were ordered based on Social Responsiveness Scale (SRS) changes between post-intervention/ follow-up and pre-intervention in SSGT and standard care groups. VCS: variant of clinical significance, VUS: variant of uncertain significance, EV: exome variant, high PRS carrier: individual who has the 10% highest PRS for ASD in our study cohort.

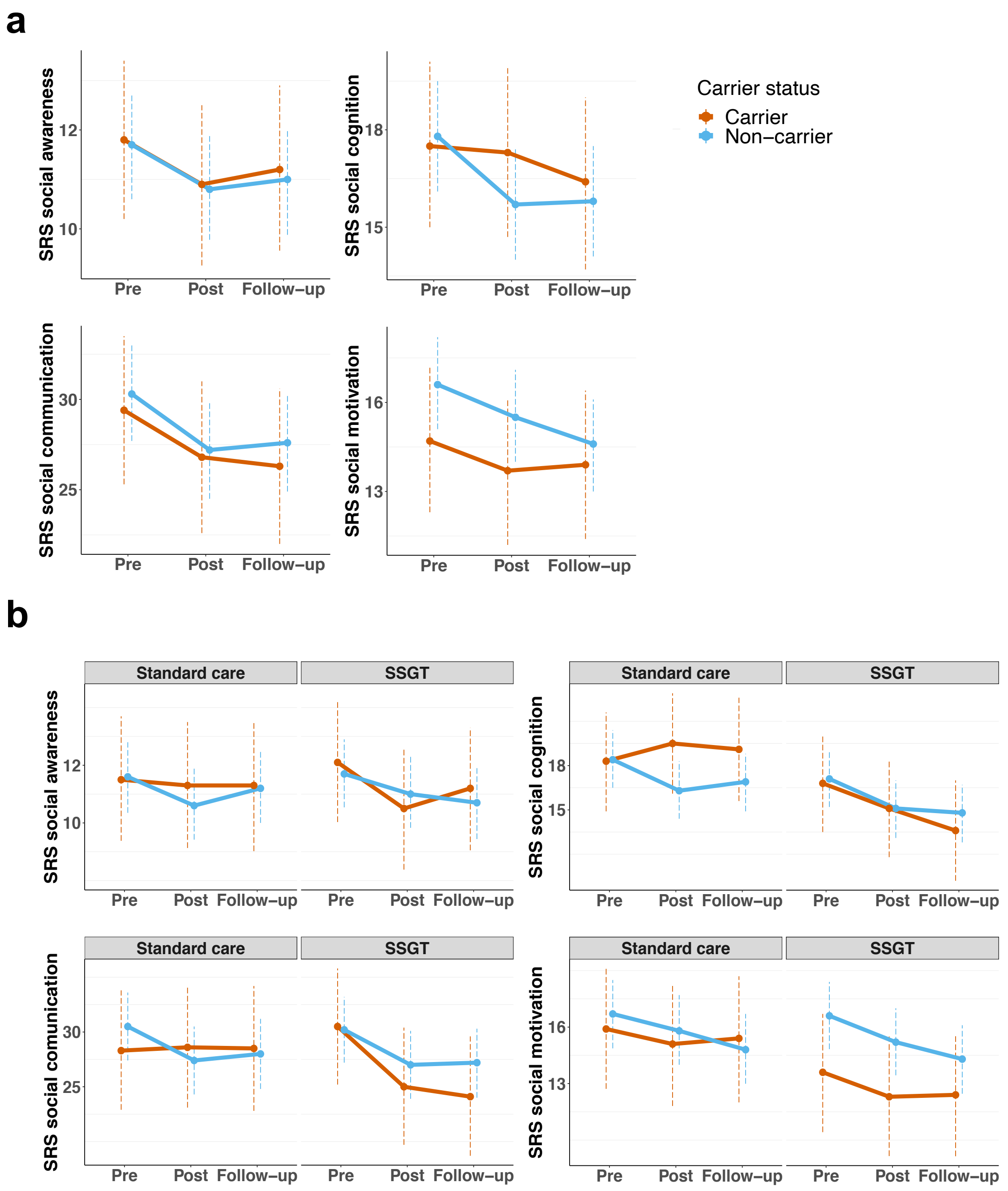

**Supplementary Figure 5**. Association between VCS+VUS carrier and intervention outcomes measured by different subscales of Social Responsiveness Scale (SRS). **a**. Least-Squares Means of SRS subscales among different variant carriers at pre-, post-intervention and follow-up. **b**. Least-Squares Means of SRS subscales among different variant carriers at pre-, post-intervention and follow-up in standard care and social skills group training (SSGT) groups. VCS: variant of clinical significance; VUS: variant of uncertain significance

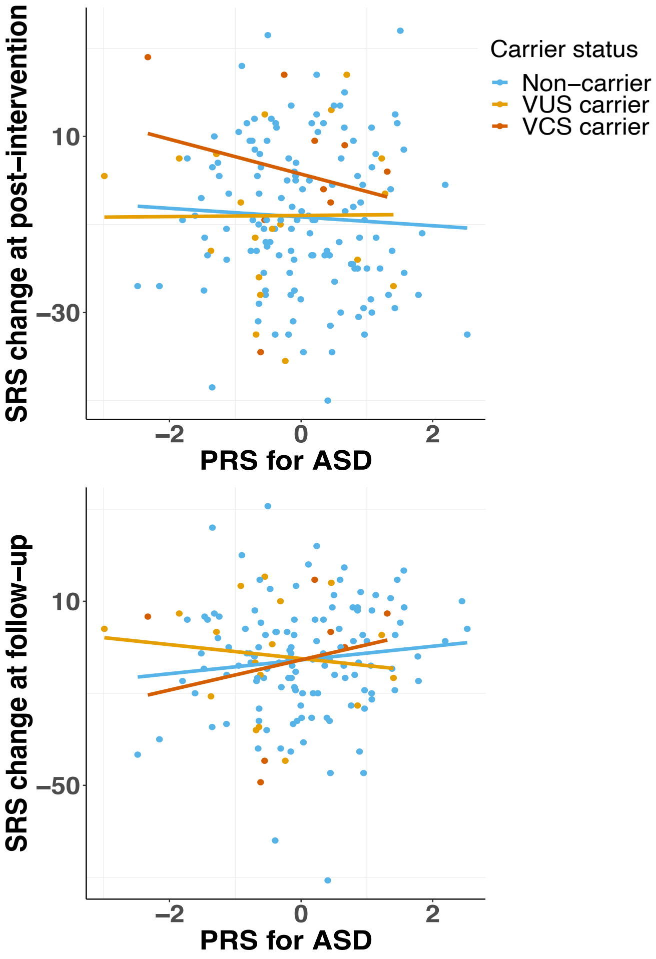

**Supplementary Figure 6.** Interaction between exome VCS/VUS and polygenic risk score (PRS) for autism spectrum disorder (ASD) at post-intervention and follow-up. Each line represents the linear model fitted by Social Responsiveness Scale (SRS) change and PRS for ASD grouped by VCS/VUS carrier status. VCS: variant of clinical significance; VUS: variant of uncertain significance

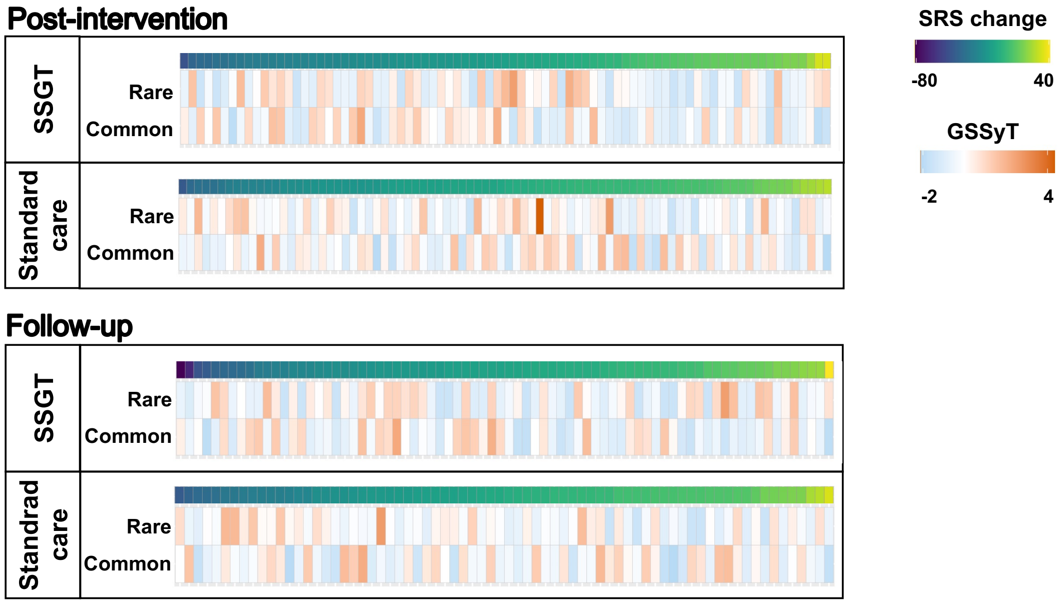

**Supplementary Figure 7**. Individual distribution of rare and common variants genetic scores for synaptic transmission genes (GSSyT) in all samples. Samples were ordered based on Social Responsiveness Scale (SRS) changes between post-intervention/follow-up and pre-intervention in social skills group training (SSGT) and standard care groups.

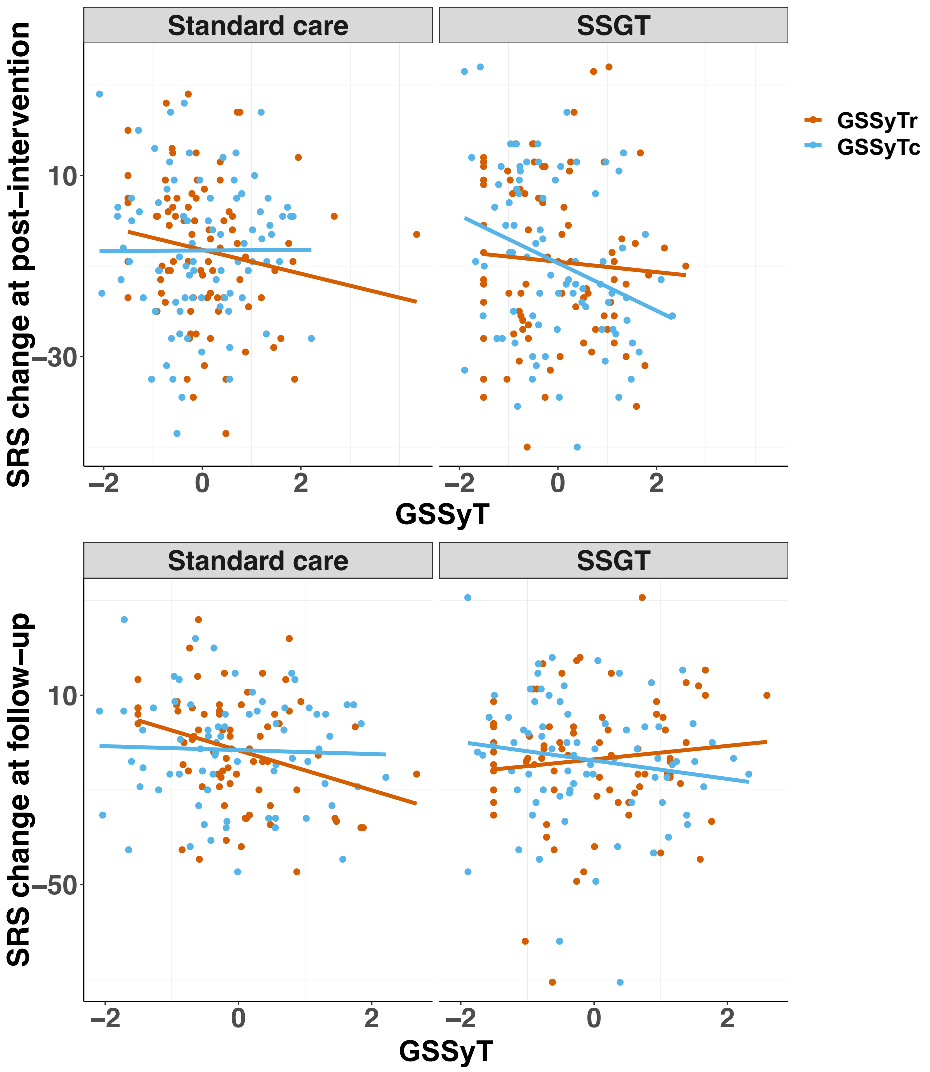

**Supplementary Figure 8**. Interaction between common and rare variants genetic scores for synaptic transmission genes ($GSSyT_c$ and $GSSyT_r$) in standard care and social skills group training (SSGT) groups. Outcomes were measured by Social Responsiveness Scale (SRS) changes between post-intervention/follow-up and pre-intervention. Each line represents the linear model fitted by SRS change and GSSyT of common or rare variants.

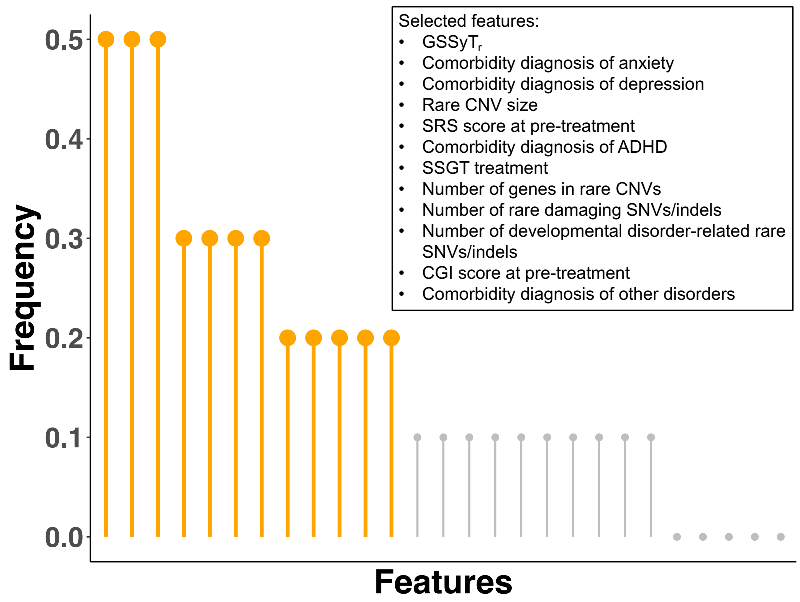

**Supplementary Figure 9**. Importance of each feature in machine learning model. $GSSyT_r$: genetic score of rare variants for synaptic transmission genes; CNV: copy number variation; SRS: Social Responsiveness Scale; ADHD: attention deficit hyperactivity disorder; SSGT: social skills group training; SNV: single nucleotide variation; CGI: Clinical Global Impression.

**Supplementary Tables**

**Supplementary Table 2**. All features used in machine learning model.

| Feature name | Type |
|---|---|
| Sample characteristics | |
| IQ | Integer |
| Age | Integer |
| Sex | Binary (0 = female, 1 = male) |
| Comorbidities | |
| Attention deficit hyperactivity disorder (ADHD) | Binary (0 = no, 1 = yes) |
| Depression | Binary (0 = no, 1 = yes) |
| Anxiety | Binary (0 = no, 1 = yes) |
| Others | Binary (0 = no, 1 = yes) |
| Treatments | |
| Social skills group training (SSGT) | Binary (0 = no, 1 = yes) |
| Cognitive behavioral therapy (CBT) | Binary (0 = no, 1 = yes) |
| Pharmaceutical treatment | Binary (0 = no, 1 = yes) |
| Counsel treatment | Binary (0 = no, 1 = yes) |
| Scales | |
| Social Responsiveness Scale (SRS) | Integer |
| Autism Diagnostic Observation Schedule (ADOS) | Integer |
| Developmental Disabilities Modification of Children's Global Assessment Scale (DD-CGAS) | Integer |
| Adaptive Behavior Assessment System (ABAS) | Integer |
| Clinical Global Impression (CGI) | Integer |
| Genetics | |
| Rare copy number variation (CNV) size | Integer |
| Number of genes in rare CNVs | Integer |
| Carrier status of clinically significant rare CNVs | Binary (0 = non-carrier, 1 = carrier) |
| Carrier status of large size rare CNVs | Binary (0 = non-carrier, 1 = carrier) |
| Polygenic risk score (PRS) for ASD | Float |
| PRS for ADHD | Float |
| Carrier status of rare exome VCS/VUS | Category (0 = non-carrier, 1 = VUS, 2 = VCS) |
| Number of rare damaging SNVs/indels | Integer |

| | |
|---|---|
| Number of developmental disorder-related rare SNVs/indels | Integer |
| $GSSyT_r$ | Float |
| $GSSyT_c$ | Float |

Abbreviations:

VCS: variant of clinical significance

VUS: variant of uncertain significance

SNV: single nucleotide variation

$GSSyT_r$: genetic score of rare variants for synaptic transmission genes

$GSSyT_c$: genetic score of common variants for synaptic transmission genes

**Supplementary Table 3**. Summary of missense variants with uncertain clinical significance.

| Gene symbol | Chr | Position | Reference allele | Alternative allele | Genotype | Transcript ID | Exon | CDS position change | Protein change | Transcripts affected/Total transcripts |
|---|---|---|---|---|---|---|---|---|---|---|
| **ACTL6B** | 7 | 100245132 | G | T | Het | ENST00000160382 | 8/14 | c.G694T | p.P232T | 1/2 |
| **CACNA1G** | 17 | 48647135 | G | A | Het | ENST00000352832 | 4/38 | c.G557A | p.R186Q | 27/28 |
| **CTCF** | 16 | 67660485 | A | G | Het | ENST00000264010 | 8/12 | c.A1385G | p.Y462C | 2/2 |
| **CUL3** | 2 | 225346771 | G | T | Het | ENST00000264414 | 14/16 | c.G1867T | p.P623T | 4/6 |
| **GRIN2B** | 12 | 13720059 | A | G | Het | ENST00000609686 | 12/13 | c.A2498G | p.L833P | 1/1 |
| **KCNH1** | 1 | 210948814 | G | A | Het | ENST00000367007 | 10/11 | c.G1988A | p.A663V | 2/2 |
| **MTOR** | 1 | 11298606 | G | A | Het | ENST00000361445 | 12/58 | c.G1855A | p.R619C | 1/3 |
| **SCN3A** | 2 | 165969493 | A | T | Het | ENST00000283254 | 21/28 | c.A3745T | p.Y1249N | 4/6 |
| **SCN8A** | 12 | 52180588 | G | A | Het | ENST00000354534 | 22/27 | c.G4205A | p.G1402E | 3/5 |
| **SMARCA2** | 9 | 2123752 | C | T | Het | ENST00000382203 | 27/34 | c.C3796T | p.R1266W | 4/16 |
| **TRIP12** | 2 | 230632446 | G | A | Het | ENST00000283943 | 41/41 | c.G5803A | p.P1935S | 3/11 |
| **TRRAP** | 7 | 98515066 | G | C | Het | ENST00000359863 | 20/72 | c.G2386C | p.G796R | 4/5 |
| **ZBTB20** | 3 | 114070690 | G | A | Het | ENST00000462705 | 9/10 | c.G16A | p.H6Y | 8/9 |
| **SYNGAP1** | 6 | 33406017 | G | C | Het | ENST00000418600 | 8/19 | c.G1335C | p.E445D | 4/4 |
| **SYNGAP1** | 6 | 33411583 | G | T | Het | ENST00000418600 | 15/19 | c.G3254T | p.R1085L | 4/4 |
| **EFTUD2** | 17 | 42930952 | G | C | Het | ENST00000426333 | 24/28 | c.G2399C | p.P800R | 6/7 |
| **NCOR1** | 17 | 15938144 | T | C | Het | ENST00000268712 | 45/46 | c.T7070C | p.H2357R | 5/14 |
| **NCKAP1** | 2 | 183850936 | C | T | Het | ENST00000361354 | 10/31 | c.C967T | p.D323N | 2/2 |
| **WHSC1** | 4 | 1902896 | G | A | Het | ENST00000503128 | 2/10 | c.G515A | p.S172N | 11/14 |
| **CHD2** | 15 | 93467702 | G | T | Het | ENST00000394196 | 3/39 | c.G214T | p.G72C | 5/7 |
| **CUX1** | 7 | 101842097 | G | T | Het | ENST00000360264 | 16/24 | c.G1910T | p.S637I | 3/12 |
| **GRIA2** | 4 | 158238831 | G | A | Het | ENST00000296526 | 5/16 | c.G688A | p.V230I | 6/11 |
| **KMT2A** | 11 | 118307595 | G | A | Het | ENST00000534358 | 1/36 | c.G368A | p.G123D | 8/9 |

| | | | | | | | | | | |
|---|---|---|---|---|---|---|---|---|---|---|
| **SHANK1** | 19 | 51190043 | G | C | Het | ENST00000293441 | 19/23 | c.G2416C | p.P806A | 4/4 |

All coordinates were based on human reference genome build GRCh37/ hg19. The transcripts were based on Ensembl GRCh37

Abbreviations:

Chr: chromosome

CDS: coding sequence

Het: heterogeneous

**Supplementary Table 4**. Association between clinically/uncertain significant variants and intervention outcomes in the whole cohort.

| | Beta | lower CI | upper CI | P |
|---|---|---|---|---|
| VCS + VUS | | | | |
| *SSGT | -1.45 | -21.41 | 18.52 | 0.89 |
| *Post-intervention | 9.22 | -0.25 | 18.70 | 0.057 |
| *Follow-up | 7.56 | -2.68 | 17.80 | 0.15 |
| *SSGT*Post-intervention | -12.16 | -25.98 | 1.66 | 0.086 |
| *SSGT*Follow-up | -10.92 | -25.47 | 3.63 | 0.14 |
| VCS | | | | |
| *SSGT | 3.85 | -36.88 | 44.59 | 0.85 |
| *Post-intervention | 14.84 | 1.60 | 28.09 | **0.029**[#] |
| *Follow-up | 6.27 | -8.67 | 21.20 | 0.41 |
| *SSGT*Post-intervention | -21.51 | -48.89 | 5.88 | 0.12 |
| *SSGT*Follow-up | -22.77 | -51.02 | 5.48 | 0.12 |
| VUS | | | | |
| *SSGT | -3.92 | -27.37 | 19.53 | 0.74 |
| *Post-intervention | 4.36 | -8.04 | 16.77 | 0.49 |
| *Follow-up | 8.09 | -4.95 | 21.12 | 0.22 |
| *SSGT*Post-intervention | -6.63 | -23.08 | 9.81 | 0.43 |
| *SSGT*Follow-up | -8.86 | -26.03 | 8.30 | 0.31 |

Abbreviations:

CI: confidence interval. [#]: $P < 0.05$

VCS: variant of clinical significance

VUS: variant of uncertain significance

**Supplementary Table 5**. Association between clinically significant variants and intervention outcome in standard care and social skills group training (SSGT) subgroups.

| | Beta | lower CI | upper CI | P |
|---|---|---|---|---|
| SSGT group | | | | |
| (VCS+VUS)*Post-intervention | -2.50 | -13.34 | 8.35 | 0.65 |
| (VCS+VUS)*Follow-up | -2.92 | -14.077 | 8.24 | 0.61 |
| VCS*Post-intervention | -6.53 | -32.46 | 19.40 | 0.62 |
| VCS*Follow-up | -16.37 | -42.32 | 9.58 | 0.22 |
| VUS* Post-intervention | -1.79 | -13.45 | 9.88 | 0.76 |
| VUS*Follow-up | -0.29 | -12.35 | 11.77 | 0.96 |
| Standard care group | | | | |
| (VCS+VUS)*Post-intervention | 9.35 | 0.70 | 18.00 | **0.036**# |
| (VCS+VUS)*Follow-up | 7.56 | -1.80 | 16.92 | 0.12 |
| VCS*Post-intervention | 14.86 | 2.79 | 26.93 | **0.017**# |
| VCS*Follow-up | 6.10 | -7.51 | 19.72 | 0.38 |
| VUS*Post-intervention | 4.60 | -6.71 | 15.91 | 0.43 |
| VUS*Follow-up | 8.21 | -3.68 | 20.10 | 0.18 |

Abbreviations:

CI: confidence interval. #: $P < 0.05$

VCS: variant of clinical significance

VUS: variant of uncertain significance

**Supplementary Table 6**. Association between genetic scores for synaptic transmission genes (GSSyT) and social skills group training (SSGT) outcome using mixed linear model.

| | Beta | lower CI | upper CI | P |
|---|---|---|---|---|
| Rare variants ($GSSyT_r$) | | | | |
| *SSGT | 3.51 | -3.68 | 10.71 | 0.34 |
| *Post-intervention | -2.73 | -6.38 | 0.92 | 0.14 |
| *Follow-up | -5.38 | -9.74 | -1.02 | **0.016**# |
| *SSGT*Post-intervention | 2.21 | -2.82 | 7.24 | 0.39 |
| *SSGT*Follow-up | 8.30 | 2.64 | 13.97 | **0.0044**# |
| Common variants ($GSSyT_c$) | | | | |
| *SSGT | 8.92 | 1.56 | 16.29 | **0.018**# |
| *Post-intervention | 0.17 | -3.40 | 3.75 | 0.92 |
| *Follow-up | -0.13 | -3.83 | 3.57 | 0.95 |
| *SSGT*Post-intervention | -5.52 | -10.59 | -0.46 | **0.033**# |
| *SSGT*Follow-up | -3.21 | -8.41 | 1.99 | 0.23 |

Abbreviation:

CI: confidence interval. #: $P < 0.05$

**Supplementary Table 7**. Association between rare/common variants genetic scores for synaptic transmission genes ($GSSyT_r$/$GSSyT_c$) and intervention outcome in standard care and social skills group training (SSGT) subgroups

| | Beta | lower CI | upper CI | P |
|---|---|---|---|---|
| SSGT group | | | | |
| $GSSyT_r$*Post-intervention | -0.51 | -4.21 | 3.18 | 0.79 |
| $GSSyT_r$*Follow-up | 2.93 | -0.93 | 6.79 | 0.14 |
| $GSSyT_c$* Post-intervention | -5.42 | -9.25 | -1.59 | **0.0062**[#] |
| $GSSyT_c$*Follow-up | -3.42 | -7.32 | 0.48 | 0.087 |
| Standard care group | | | | |
| $GSSyT_r$*Post-intervention | -2.74 | -6.12 | 0.65 | 0.11 |
| $GSSyT_r$*Follow-up | -5.46 | -9.50 | -1.41 | **0.0091**[#] |
| $GSSyT_c$*Post-intervention | 0.16 | -3.15 | 3.47 | 0.93 |
| $GSSyT_c$*Follow-up | -0.16 | -3.58 | 3.27 | 0.93 |

Abbreviation:

CI: confidence interval. [#]: $P < 0.05$
